# SARS-CoV-2 mRNA vaccination exposes progressive adaptive immune dysfunction in patients with chronic lymphocytic leukemia

**DOI:** 10.1101/2022.12.19.22283645

**Authors:** Kai Qin, Kazuhito Honjo, Scott Sherrill-Mix, Weimin Liu, Regina Stoltz, Allisa K. Oman, Lucinda A. Hall, Ran Li, Sarah Sterrett, Ellen R. Frederick, Jeffrey R. Lancaster, Mayur Narkhede, Amitkumar Mehta, Foluso J. Ogunsile, Rima B. Patel, Thomas J. Ketas, Victor M Cruz Portillo, Albert Cupo, Benjamin M. Larimer, Anju Bansal, Paul A. Goepfert, Beatrice H. Hahn, Randall S. Davis

**Affiliations:** Department of Medicine, University of Alabama at Birmingham, Birmingham, AL 35294, USA; Department of Medicine, University of Pennsylvania, Philadelphia, PA 19104, USA; Department of Radiology, University of Alabama at Birmingham, Birmingham, AL 35294, USA; O’Neal Comprehensive Cancer Center, University of Alabama at Birmingham, Birmingham, AL 35294, USA; Department of Microbiology and Immunology, Weill Medical College of Cornell University, New York, NY 10065, USA; Department of Microbiology, University of Alabama at Birmingham, Birmingham, AL 35294, USA; Department of Microbiology, University of Pennsylvania, Philadelphia, PA 19104, USA; Department of Biochemistry & Molecular Genetics, University of Alabama at Birmingham, Birmingham, AL 35294, USA

## Abstract

Chronic lymphocytic leukemia (CLL) patients have lower seroconversion rates and antibody titers following SARS-CoV-2 vaccination, but the reasons for this diminished response are poorly understood. Here, we studied humoral and cellular responses in 95 CLL patients and 30 healthy controls after two BNT162b2 or mRNA-2173 mRNA immunizations. We found that 42% of CLL vaccinees developed SARS-CoV-2-specific binding and neutralizing antibodies (NAbs), while 32% had no response. Interestingly, 26% were seropositive, but had no detectable NAbs, suggesting the maintenance of pre-existing endemic human coronavirus-specific antibodies that cross-react with the S2 domain of the SARS-CoV-2 spike. These individuals had more advanced disease. In treatment-naïve CLL patients, mRNA-2173 induced 12-fold higher NAb titers and 1.7-fold higher response rates than BNT162b2. These data reveal a graded loss of immune function, with pre-existing memory being preserved longer than the capacity to respond to new antigens, and identify mRNA-2173 as a superior vaccine for CLL patients.

## INTRODUCTION

Chronic lymphocytic leukemia (CLL) is the most prevalent leukemia in Western countries and mainly affects the elderly, with a median age at diagnosis of 70 years.^1^ Because the natural progression of this B cell malignancy as well as its treatments weaken adaptive and innate immunity, infections are a leading cause of death.^2^ Most patients are followed with a ‘watch and wait’ strategy for years until they meet criteria for therapy.^3^ However, vaccine responses even in treatment-naïve patients are impaired^4–8^ and many require intravenous immunoglobulin (IVIg) infusions to mitigate infections.^9^ The mechanisms responsible for the loss of immune function are still poorly understood.

Since their emergence hundreds of years ago,^10^ four human coronaviruses (HCoV-229E, HCoV-NL63, HCoV-HKU1, and HCoV-OC43), which cause mild seasonal upper respiratory infections,^11^ have become endemic. However, more recently there have been zoonotic outbreaks of three pathogenic HCoVs, including severe acute respiratory syndrome-coronavirus (SARS)-CoV, Middle East Respiratory Syndrome (MERS)-CoV, and most recently SARS-CoV-2. The coronavirus disease of 2019 (COVID-19) pandemic caused by SARS-CoV-2 has resulted in the deaths of over 6 million people globally and over 1 million in the United States.^12^ Severe illness and mortality are especially high in older individuals with comorbidities and compromised immunity.^13, 14^ Hence, the SARS-CoV-2 pandemic has posed a particularly difficult challenge for CLL patients, which is underscored by two international studies that demonstrated COVID-19 fatality rates of ∼27-38%.^15, 16^ Although mortality rates have decreased with mitigation strategies and the evolution of less pathogenic variants,^17^ preventing SARS-CoV-2 infection in these individuals remains a high priority.

SARS-CoV-2 enters human respiratory epithelial cells following the binding of the viral spike (S) glycoprotein receptor binding domain (RBD), located within the S1 subunit, with the host angiotensin-converting enzyme 2 (ACE2) receptor.^18, 19^ Thus, neutralizing antibodies (NAbs) that disrupt RBD/ACE2 binding and viral entry represent a key defense. NAbs represent an important correlate of immune protection, as evidenced by the beneficial effects of convalescent plasma (CP) and recombinant NAbs in patients who are unable to mount an adequate antiviral response.^20–22^ In addition, NAbs prevent symptomatic SARS-CoV-2 infection in immunocompetent hosts.^23^

Multiple reports indicate diminished immune responses in CLL patients following COVID-19 mRNA vaccination,^24–26^ but the reasons for this decreased reactivity even in treatment-naïve patients remain largely unknown. Here we studied humoral and cellular immune responses in a clinically well-characterized cohort of SARS-CoV-2 infection-naïve CLL patients and healthy controls following two immunizations with the Pfizer-BioNTech (BNT162b2) or Moderna (mRNA-2173) mRNA vaccines. Consistent with previous reports, we show that both cellular and humoral vaccine-induced responses are reduced in CLL patients. Moreover, vaccinees exhibited a wide variety of antibody responses, ranging from only moderately diminished binding and NAb titers to a complete absence of detectable antibodies. One group of vaccinees was of particular interest since they failed to develop SARS-CoV-2 specific NAbs, but had high-titer binding antibodies that preferentially reacted with the S2 subunit of the SARS-CoV-2 spike. Since these individuals also exhibited high-titer seroreactivity to endemic HCoVs, their anti-HCoV antibodies likely cross-reacted with conserved regions of the SARS-CoV-2 spike. Thus, SARS-CoV-2 vaccination exposed a graded decline in immune function, with a subset of CLL patients still being able to maintain, and possibly boost, pre-existing immune responses, while having lost the ability to respond to new antigens.

## RESULTS

### Study participants

We recruited 95 patients diagnosed with CLL according to IWCLL criteria^3^ and 30 healthy controls with no prior SARS-CoV-2 infection as determined by a lack of nucleocapsid antibodies. All participants received two doses of either the Pfizer BNT162b2 or the Moderna mRNA-2173 vaccine, both of which encode the SARS-CoV-2 Wuhan 1 spike protein. Clinical characteristics of the enrolled CLL patients and healthy controls are provided in Tables 1 and S1. The median age of the CLL patients was 72 years (IQR, 64-77) and 47 were male. Forty-five of the patients (47%) were treatment-naïve, whereas 50 (53%) had prior therapy, including 34 who were actively treated (i.e., anti-CD20 therapy, Bruton’s tyrosine kinase [BTK] inhibition). Seven individuals were refractory to therapy and relapsed, and nine were off-therapy in clinical remission. Sixty-one patients (64%) received 30 µg of the Pfizer BNT162b2 vaccine, while 34 (36%) were immunized with 100 µg of the Moderna mRNA-2173 vaccine according to FDA guidelines. The median time period from the second immunization to testing was 38 days (IQR, 26-83) for CLL donors and 35 days (IQR, 28-56.5) for healthy controls.

**Table 1.** Clinical and disease characteristics of healthy controls and CLL patients.

| Characteristic | Healthy controls<br>(n=30) | CLL<br>(n=95) |
| --- | --- | --- |
| Age in years, median (IQR) | 62 (44-69.8) | 72 (64-77) |
| Age ≥65 y, n (%) | 13 (43.3) | 71 (74.7) |
| Sex, male, n (%) | 16 (53.3) | 47 (49.5) |
| **Rai Stage\*, n (%)** |  |  |
| 0-I | - | 46 (88.5) |
| II-IV | - | 6 (11.5) |
| **Disease/Treatment status, n (%)** |  |  |
| Treatment-naïve | - | 45 (47.4) |
| Active-therapy | - | 34 (35.8) |
| Off-therapy in remission | - | 9 (9.5) |
| Off-therapy in relapse | - | 7 (7.4) |
| **Molecular and phenotypic biomarkers** |  |  |
| *IGHV*, mutated, n (%) | - | 47/75 (62.7) |
| CD38 (≥20%), n (%) | - | 23/90 (25.6) |
| **FISH, n (%)** |  |  |
| Normal | - | 10 (11.1) |
| del(13q) | - | 48 (53.3) |
| Trisomy 12 | - | 15 (16.7) |
| del(11q) | - | 10 (11.1) |
| del(17p) | - | 7 (7.8) |
| **IVIg therapy, n (%)** | - | 25 (26.3) |
| **Laboratory parameters, median (IQR)** |  |  |
| Absolute lymphocyte count, (10<sup>9</sup>/L) | - | 6.8 (1.9-21.4) |
| β2-microglobulin, mg/L | - | 2.2 (1.8-3.1) |
| IgM, mg/dL | - | 28 (19-62.5) |
| IgG, mg/dL | - | 713 (546.5-974) |
| IgA, mg/dL | - | 105 (63.8-162.8) |

IGHV, immune globulin heavy chain variable gene; FISH, fluorescence in situ hybridization
\*Determined for treatment-naïve and patients off therapy in relapse

### Binding and neutralizing antibody responses in CLL patients correlate with disease status

Plasma samples from all CLL vaccinees and healthy controls were tested for SARS-CoV-2 anti-spike and RBD binding antibodies by ELISA as described.^22, 27^ While all control subjects generated both anti-S and RBD IgG following immunization, response rates were significantly reduced in CLL patients, with only 65 (68%) developing anti-S and 51 (54%) developing RBD antibodies (Tables S2 and S3). CLL vaccinees also had 23-fold lower anti-S and 30-fold lower RBD half-maximal effective concentrations (EC_50_) compared to healthy controls (Figures 1A and 1B). Even when comparing only CLL patients who mounted a humoral response (i.e., CLL responders), we found median IgG anti-S and RBD EC_50_ values that were 7.2-fold and 6.4-fold lower than those of healthy controls, respectively (Tables S2 and S3).

We next explored whether there was an association between seroreactivity and disease history. As expected, treatment-naïve patients had higher response rates and anti-S Ab titers (median 2,733) compared to vaccinees on active CLL therapy (median <100) (Figure 1C). All CLL patients who were in clinical remission (CR) mounted anti-S responses and had significantly higher IgG titers (median 2,740) compared to those on treatment. RBD response rates and Ab titers were also higher for individuals who were treatment-naive or in clinical remission compared to actively treated patients, and vaccinees who were refractory to therapy or relapsed (R/R) had generally lower anti-S and RBD responses (Figures 1C and 1D). Although most individuals who were treatment-naive or in clinical remission had anti-S and RBD IgG binding antibodies, their titers were significantly lower compared to controls (Figures 1A-1D).

**Figure 1.**
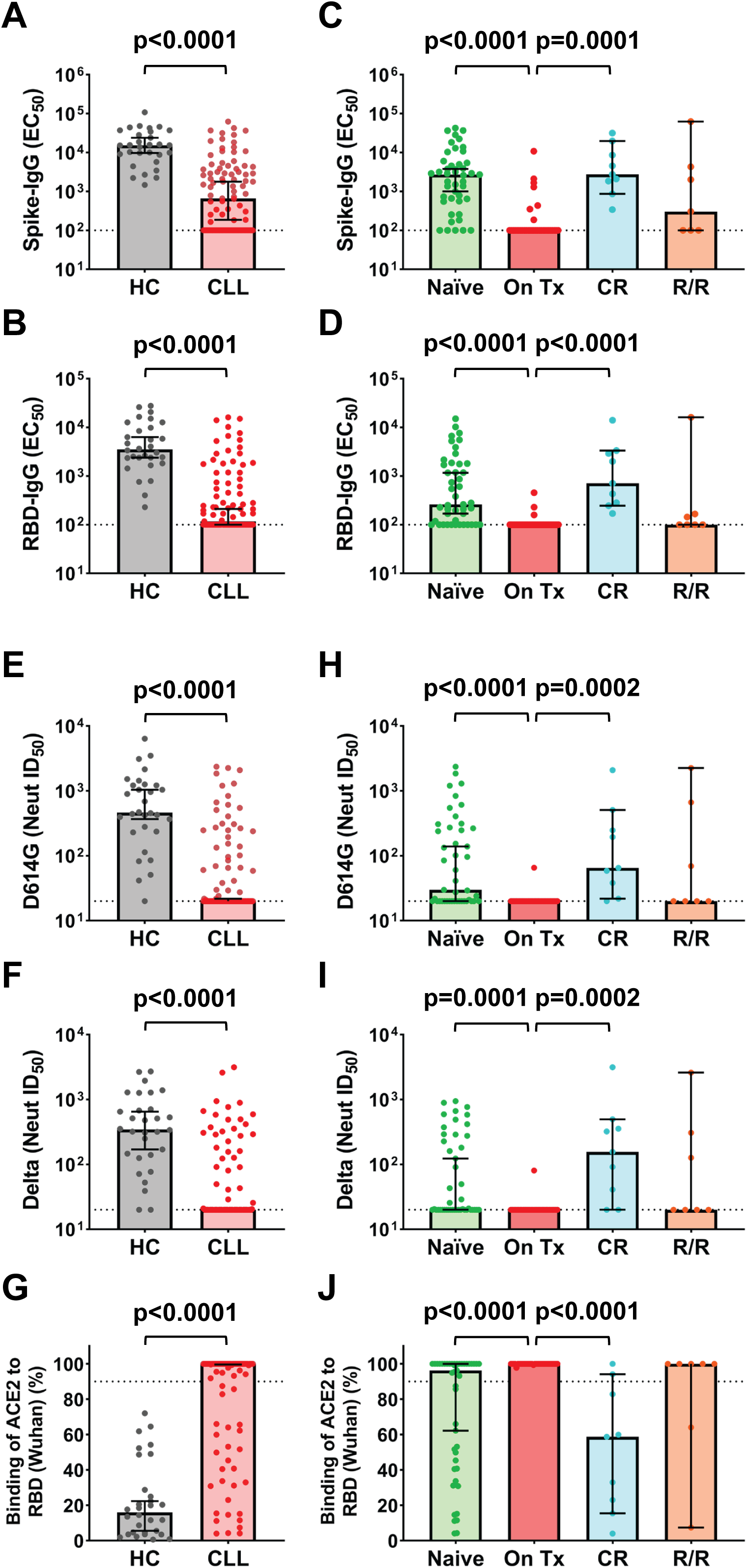
Serologic and neutralizing responses in SARS-CoV-2 mRNA vaccinated CLL patients and healthy controls (HC). (A and B) Comparison of SARS-CoV-2 vaccine-elicited spike (A) and RBD (B) ELISA IgG antibody titers in HC (n=30) and CLL (n=95) patients expressed as half-maximal effective concentrations (EC_50_). (C and D) SARS-CoV-2 vaccine-elicited spike (C) and RBD (D) ELISA IgG antibody titers stratified by CLL disease status: treatment-naïve (Naïve) (n=45), on-therapy (On Tx) (n=34), off-therapy in clinical remission (CR) (n=9), and off therapy and relapsed or refractory (R/R) (n=7). (E-G) Comparison of SARS-CoV-2 vaccine-elicited neutralizing antibody titers for HC (n=30) and CLL patients against (E) D614G (CLL; n=95) and (F) delta (CLL; n=93) spike variants expressed as the reciprocal half-maximal inhibitory dilution (ID_50_) as determined in an HIV-based pseudovirus neutralization assay or by (G) ACE2/RBD (Wuhan) binding inhibition (CLL; n=95) at a 1:25 dilution. (H-J) SARS-CoV-2 vaccine elicited NAb titers for D614G (H) and delta (I) S variants and ACE2/RBD binding frequencies (J) stratified by CLL disease status. Bars indicate the median with 95% CI. Dotted black lines indicate assay sensitivity cutoffs (EC_50_ values of <100 by ELISA, ID_50_ values of <20 in the neutralization assay, and >90% ACE2 binding in the RBD-inhibition assay). P values were determined by the Mann-Whitney test (A-B, E-G) or Dunn’s test of multiple comparisons following a Kruskal-Wallis test (C-D, H-J).

We next analyzed plasma samples for neutralizing activity against the SARS-CoV-2 D614G^28^ and B.1.617.2 (also known as delta)^29, 30^ variants using an HIV-1-based pseudovirus assay.^31^ Consistent with the ELISA findings, neutralizing responses in CLL patients were overall reduced. While NAbs against the D614G and delta variants were found in 97% and 93% of healthy controls, respectively, NAb response rates in CLL patients were significantly lower at 42% (40/95) and 38% (35/93) (Tables S2 and S3). An assay that measured Ab-mediated inhibition of the ACE2/RBD (Wuhan strain) interaction,^22^ yielded very similar results, detecting blockade in all controls, but in only 30% (28/95) of CLL patients (Tables S2 and S3). Median NAb titers were also significantly lower in CLL patients than healthy controls, with half-maximal inhibitory dilutions (ID_50_) for D614G being >23-fold (464 vs ≤ 20; Figure 1E) and Delta being >17-fold (346 vs ≤ 20; Figure 1F) lower, respectively. Similarly, ACE2/RBD inhibition was lower in CLL patients (Figure 1G). Finally, neutralizing responses in CLL patients reflected their disease and treatment status. Response rates, NAb titers, and ACE2/RBD blockade were all significantly higher in individuals who were treatment-naive or in clinical remission compared to individuals on active treatment or in relapse (Figures 1H-1J). These data confirm and extend earlier findings,^24, 25, 32^ showing impaired humoral responses not only in treated but also in treatment-naïve CLL patients.

### Clinical predictors of humoral immune responses in vaccinated CLL patients

To search for predictors of humoral responses following immunization, we analyzed the demographics and disease characteristics of CLL vaccinees. By univariate analysis we compared 18 clinical variables with binding (anti-S and RBD IgG) and neutralizing (D614G, Delta, and ACE2/RBD blockade) antibody responses, measured as binary parameters (Tables 1, S1, and S4). Examining the four CLL groups (treatment-naive, active therapy, clinical remission, and refractory/relapsed), disease status itself was significantly associated with responsivity (Table S4A). Clinical determinants that correlated with higher IgG titers and neutralizing activity included early Rai stage disease, low serum β2-microglobulin (≤2.4 mg/L) levels, lack of prior CLL therapy, vaccination ≥ 12 months following anti-CD20 therapy, and no requirement for IVIg therapy. Since D614G and Delta NAb titers were largely equivalent, to determine clinical risk factors associated with a failure to mount anti-S IgG and/or NAb responses, we performed a multivariate analysis using only the D614G NAb data (Table S4B). As expected, active therapy was a significant adverse predictor of both anti-S binding (OR, 62; 95% CI, 3.6-3500) and D614G NAbs (OR, 40; 95% CI, 1.2-2500), whereas being refractory to therapy (OR, 7.7; 95% CI, 1.1-64) and requiring prophylactic IVIg therapy (OR, 5.2; 95% CI, 1.3-29) were both associated with poor humoral responses. Unexpectedly, BNT162b2 vaccination was also a negative predictor of D614G NAbs (OR, 5.8; 95% CI, 1.6-27), suggesting vaccine-specific differences in neutralizing antibody elicitation.

### Vaccinated CLL patients have reduced CD4^+^ but relatively preserved CD8^+^ T cell functions

To investigate the impact of vaccination on cell-mediated immunity, we examined peripheral blood mononuclear cells (PBMCs) from a subset of vaccinated CLL patients (n=36) and healthy controls (n=21) for which sufficient samples were available (Table S5). The frequencies of circulating total CD3^+^, CD4^+^, and CD8^+^ T cells as well as naïve and memory subsets were determined by multi-color flow cytometry analysis with CD45RA and CCR7 immunophenotyping (Figure S1 and Table S5A). As expected, total CD3^+^ T cell frequencies were significantly higher in controls than CLL vaccinees (Figure S1A). Immunophenotypic analysis showed skewing of the CLL T cell compartment, with lower total CD4^+^ and higher CD8^+^ T cell numbers, resulting in lower CD4:CD8 ratios compared to controls (Figures S1B-S1D). Among T cell subsets, CLL vaccinees had relatively lower frequencies of naïve CD4^+^ and CD8^+^ T cells (Figures S1E and S1F). In contrast, CLL effector memory CD8^+^, but not CD4^+^, T cell frequencies were higher. However, central memory and terminally differentiated effector memory RA T cells did not differ between the groups. These results confirmed previous findings,^33–36^ indicating lower naïve CD4^+^ and CD8^+^ T cell populations as well as higher effector memory CD8^+^ T cells in CLL vaccinees compared to healthy controls.

To examine antigen-specific function, we determined activation-induced marker (AIM) expression for CD4^+^, circulating T follicular helper cell (cTfh), and CD8^+^ T cells following S and N peptide pool (Wuhan strain) stimulation. For both CD4^+^ T and cTfh cells, antigen specificity was quantified by the frequency of PD-L1 and OX40 co-expressing cells, while the CD69^+^CD137^+^ population was used to identify CD8^+^ T cells (Figure S2A; Tables S5B and S5C). Although all subjects lacked nucleocapsid (N) antigen seroreactivity, AIM T cell responses against N peptides were detected in the CD8^+^ T cells of one healthy control and the CD4^+^ T cells of three CLL patients, likely representing responses to prior endemic HCoV infections (Table S5B).^37, 38^ In contrast, overall responses against S peptides were found in 91% of controls, but only 33% of CLL patients (Table S5C). S-restricted responder rates for CLL vaccinees were also significantly lower for each of the three T cell subsets analyzed (Figure 2A). Moreover, the median frequencies of S reactive AIM responding cells among the three T cell subsets were significantly reduced in CLL vaccinees compared to healthy controls (Figure 2B). These findings demonstrate lower antigen-specific responses by different T cell subsets in CLL patients following SARS-CoV-2 mRNA vaccination.

**Figure 2.**
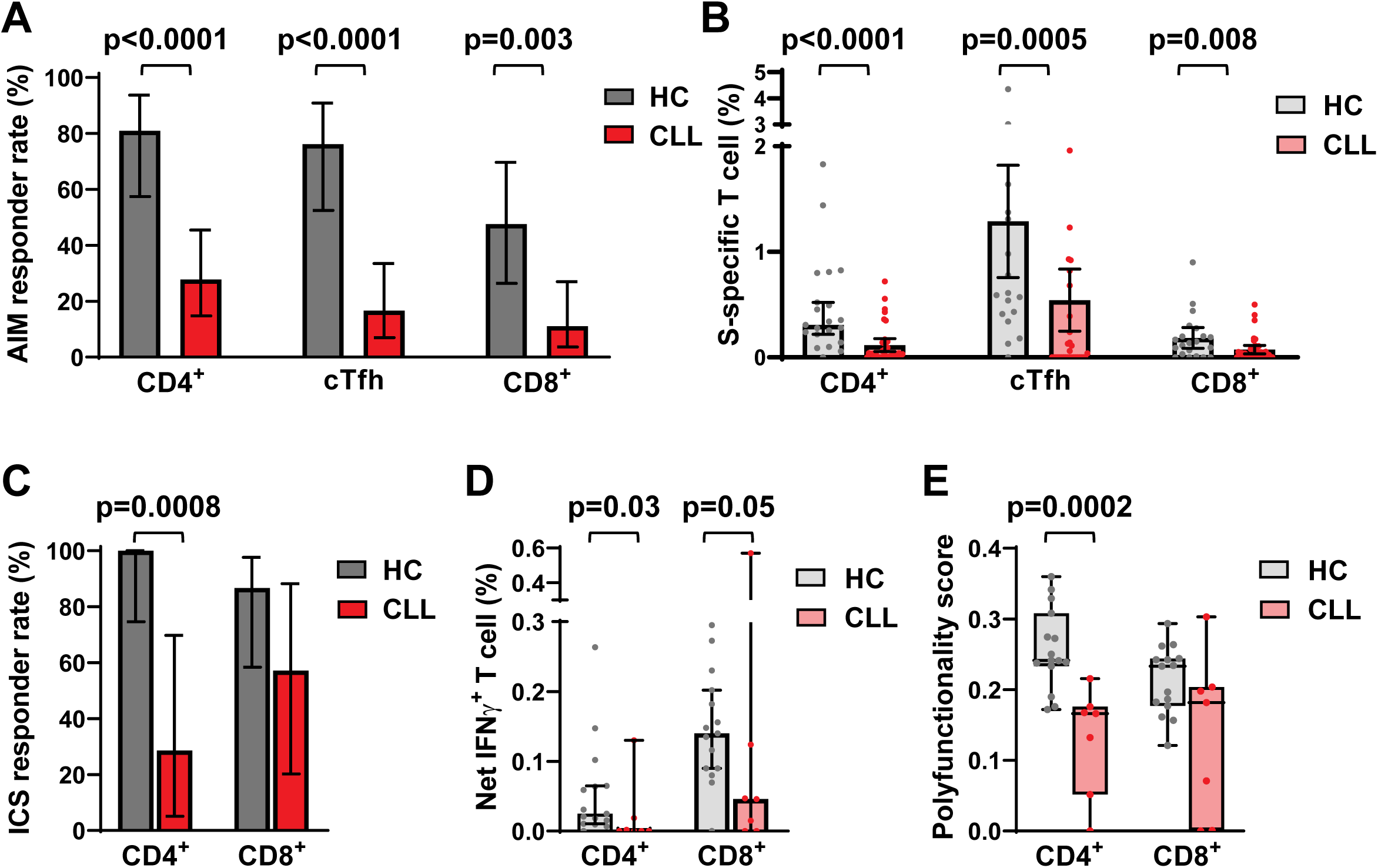
Impaired S peptide-restricted T cell subset responses and CD4^+^ effector function, but retained CD8^+^ T cell reactivity in CLL vaccinees. (A) Comparisons of S-specific T cell AIM response rates among CD4^+^, cTfh, and CD8^+^ T cell subpopulations in SARS-CoV-2 mRNA vaccinated HC and CLL patients. (B) Quantitative comparisons of S-specific T cell AIM response frequencies for CD4^+^, cTfh, and CD8^+^ T cells in SARS-CoV-2 mRNA vaccinated HC and CLL patients. (C) ICS response rates of S-specific CD4^+^ and CD8^+^ T cells from HC and CLL donors. Responders were defined as individuals with reactivity against at least one of five effector features (see text) upon peptide stimulation. (D) Quantitative comparisons of IFNγ production by CD4^+^ and CD8^+^ T cells. (E) Comparisons of CD4^+^ and CD8^+^ T cell effector responses calculated using combinatorial polyfunctionality analysis.^39^ Bars indicate the (A and C) mean or (B, D-E) median with 95% CI. P values were determined by Fisher’s exact test (A and C) or the Mann-Whitney test (B, D-E).

We next tested CD4^+^ and CD8^+^ T cell function by quantifying cytokine and effector molecule production by intra-cellular staining (ICS). To exclude potential effects of prior HCoV infections, only healthy controls (n=15) and CLL (n=7) samples with positive S and negative N peptide responses were analyzed. ICS positivity was defined by a T cell response to at least one of five parameters: IFNγ, IL-2, TNFα, CD107a plus granzyme B, or CD107a plus perforin (Figure S2B). S-restricted CD4^+^ T cell responses were significantly higher in controls compared to CLL vaccinees (Figure 2C and Table S5D), with most pronounced differences observed for IFNγ (Figure 2D). By combinatorial polyfunctionality analysis,^39^ a higher score for CD4^+^ T cells indicated more robust effector function for this subset in healthy controls compared to CLL vaccinees (Figure 2E). In contrast, the responder rate and single or polyfunctionality quantitation for CD8^+^ T cells was comparable between the cohorts, although CLL vaccinees showed a trend toward lower IFNγ production. The difference between CD4^+^ and CD8^+^ T cell function was generally consistent between the AIM and ICS analyses. Univariate analyses to define potential clinical correlates with these T cell studies failed to identify significant associations, likely reflecting the low response frequencies and testing of a subset of the total cohort. Overall, these data indicate reduced S-restricted CD4^+^ T cell effector functions, but relatively preserved CD8^+^ T cell functions in CLL vaccinees.

### Vaccinees who lack SARS-CoV-2 NAbs maintain antibodies to endemic coronaviruses

An analysis of the serologic data in Figure 1 showed that 32% of all CLL patients failed to seroconvert or develop D614G NAbs following vaccination (S^-^NAb^-^), while 42% developed both anti-S binding antibodies and NAbs (S^+^NAb^+^) (Figure 3A). Unexpectedly, 26% of CLL vaccinees had anti-S binding antibodies, but lacked detectable NAbs (S^+^NAb^-^). This was not due to IVIg treatment, since among the 25 IVIg treated CLL patients 14 were S^-^NAb^-^ and 6 were S^+^NAb^-^. Instead, the data suggested that some CLL patients had selectively lost their ability to generate NAbs despite exhibiting anti-S reactivity. To examine this unusual phenotype, we compared S and RBD binding antibody titers in S^+^NAb^+^ and S^+^NAb^-^ CLL patients. Median IgG titers against these two antigens were 3 to 4-fold lower for S^+^NAb^+^ CLL patients compared to healthy controls, but were even more diminished for individuals with S^+^NAb^-^ status, i.e., 23-fold and 35-fold, respectively (Figures 3B and 3C).

**Figure 3.**
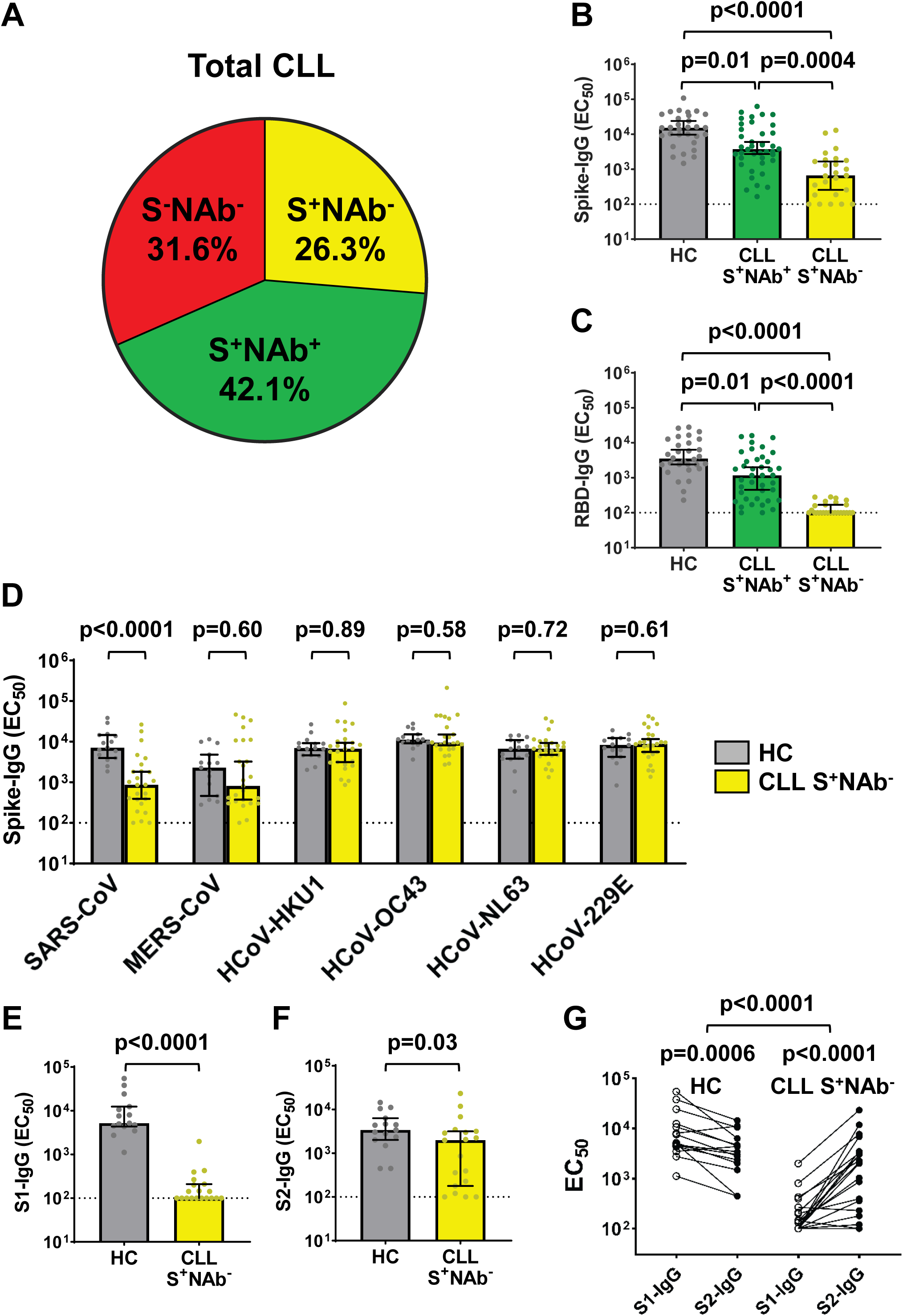
SARS-CoV-2 S-reactive CLL vaccinees lacking NAbs have pre-existing cross-reactivity with endemic HCoVs and the S2 subunit preferentially. (A) Pie chart illustrating the distribution of SARS-CoV-2 S-specific seroreactivity and NAb generation (D614G), which define three different serologic profiles among all CLL patients. (B and C) Comparisons of IgG EC_50_ titers against SARS-CoV-2 (Wuhan) full-length S (B) or RBD (C) for all HC (n=30) versus S^+^NAb^+^ (n=40) and S^+^NAb^-^ (n=25) patients defined by the detection of endpoint titer Ab reactivity quantitated by ELISA. (D) Comparisons of IgG EC_50_ titers against the S (S1+S2) proteins of two pathogenic and four endemic HCoVs measured by ELISA from SARS-CoV-2 vaccinated HC (n=15) and CLL S^+^NAb^-^ (n=25) samples. (E and F) Comparisons of IgG EC_50_ titers against SARS-CoV-2 S1 (E) or S2 (F) domains in vaccinated HC (n=15, from D) versus S^+^NAb^-^ (n=19) CLL donors. (G) Paired comparisons of log-transformed S1 versus S2 EC_50_ titers for the HC and S^+^NAb^-^ CLL donors analyzed in (E and F). Bars indicate the median (B-F) with 95% CI. Dotted black lines indicate assay sensitivity cutoffs, specifically, EC_50_ values of <100. P values were determined by Dunn’s test of multiple comparisons following a Kruskal-Wallis test (B and C), the Mann-Whitney test (D-F), and with a two-tailed paired or unpaired t-test for differences within and between cohorts (G).

One potential explanation for the S^+^NAb^-^ serologic phenotype was the presence of cross-reactive antibodies from prior HCoV infections. To investigate this possibility, we tested all S^+^NAb^-^ CLL patients (n=25) and a subset of the healthy controls (n=15) for IgG binding to recombinant spike proteins of six HCoVs: SARS-CoV, MERS-CoV, HCoV-HKU1, HCoV-OC43, HCoV-NL63, and HCoV-229E. We found that IgG EC_50_ titers against the SARS-CoV spike, which shares ∼75% ectodomain sequence identity with SARS-CoV-2,^18, 19^ were significantly lower for S^+^NAb^-^ CLL patients than controls, and a similar trend was observed for the more distantly related MERS-CoV spike. However, no such differences were found for the other HCoV spike proteins, against which high-titer antibodies were detected in both CLL vaccinees and heathy controls (Figure 3D). Thus, while S^+^NAb^-^ vaccinees were unable to produce SARS-CoV-2 specific NAbs, they maintained high-titer HCoV-specific antibodies that cross-reacted with the spike proteins of SARS-CoV and MERS-CoV.

To further dissect the S^+^NAb^-^ serotype, we analyzed available samples from S^+^NAb^-^ CLL vaccinees (n=19) and healthy controls (n=15) for IgG binding against the SARS-CoV-2 S1 and S2 subunits by ELISA. CLL EC_50_ titers against these two S proteins were again significantly lower compared to controls; however, this difference was much more pronounced for S1 (52-fold) than S2 (1.7-fold) (Figures 3E and 3F, Table S2). Moreover, a paired donor analysis of S1 and S2-IgG EC_50_ titers revealed that S^+^NAb^-^ CLL vaccinees had higher S2 titers than the controls who had elevated S1 titers (Figure 3G). This S2 bias remained significant even when median differences in S2-S1 titers were subtracted. Taken together, these data suggest that S^+^NAb^-^ vaccinees were able to maintain, and possibly even boost, pre-existing HCoV antibodies that cross-reacted with conserved epitopes in the S2 subunit of SARS-CoV-2. Unfortunately, the latter possibility could not be tested since pre-vaccination samples were not available.

### The mRNA-1273 vaccine elicits superior NAb responses in treatment-naive CLL patients

A multivariate analysis identified significantly higher D614G NAb response rates in CLL patients vaccinated with mRNA-2173 (53%, 18/34) compared to those vaccinated with BNT162b2 (36%, 22/61), despite very similar treatment and clinical states (Fisher’s exact test, p = 0.73) (Figure 4A, Table S4B). To examine the reason for this difference, we compared NAb ID_50_ titers in all patients by vaccine type. Remarkably, both the median D614G and Delta ID_50_ NAb titers of mRNA-2173 immunized CLL patients were significantly higher than those of CLL vaccinees who received BNT162b2 (Figures 4B and S3A). Because a large number of patients was unable to mount a humoral response because of CLL-directed immunosuppressive therapy (Figure 4B, red samples), we next focused on treatment-naïve BNT162b2 (n=30) and mRNA-2173 (n=15) CLL vaccinees. Again, these two groups did not differ in parameters of clinical progression, including Rai stages, absolute lymphocyte counts (ALC), serum β2M levels, and IVIg prophylaxis requirements (Figure 4C). Although only 28 of 45 treatment-naïve CLL vaccinees developed D614G NAbs, the response rates were significantly higher for mRNA-2173 (13/15, 87%) than for BNT162b2 (15/30, 50%) recipients (Figure 4D). Accordingly, mRNA-2173 vaccinees had 6.5-fold higher odds (95% CI, 1.3-31.7) of NAb development than BNT162b2 recipients. Similar results were obtained for the Delta variant, where 11 of 15 (73%) mRNA-2173 vaccinees mounted detectable NAbs compared to 13 of 30 (43%) BNT162b2 recipients, although these differences did not reach significance (Figure S3B). Finally, both D614G and Delta NAb titers were significantly higher in mRNA-2173 compared to BNT162b2 recipients (Figures 4E and S3C), despite very similar clinical characteristics (Figure 4C). Thus, the inferior NAb responses in CLL patients immunized with BNT162b2 were a consequence of the vaccine type and not differences in disease progression.

**Figure 4.**
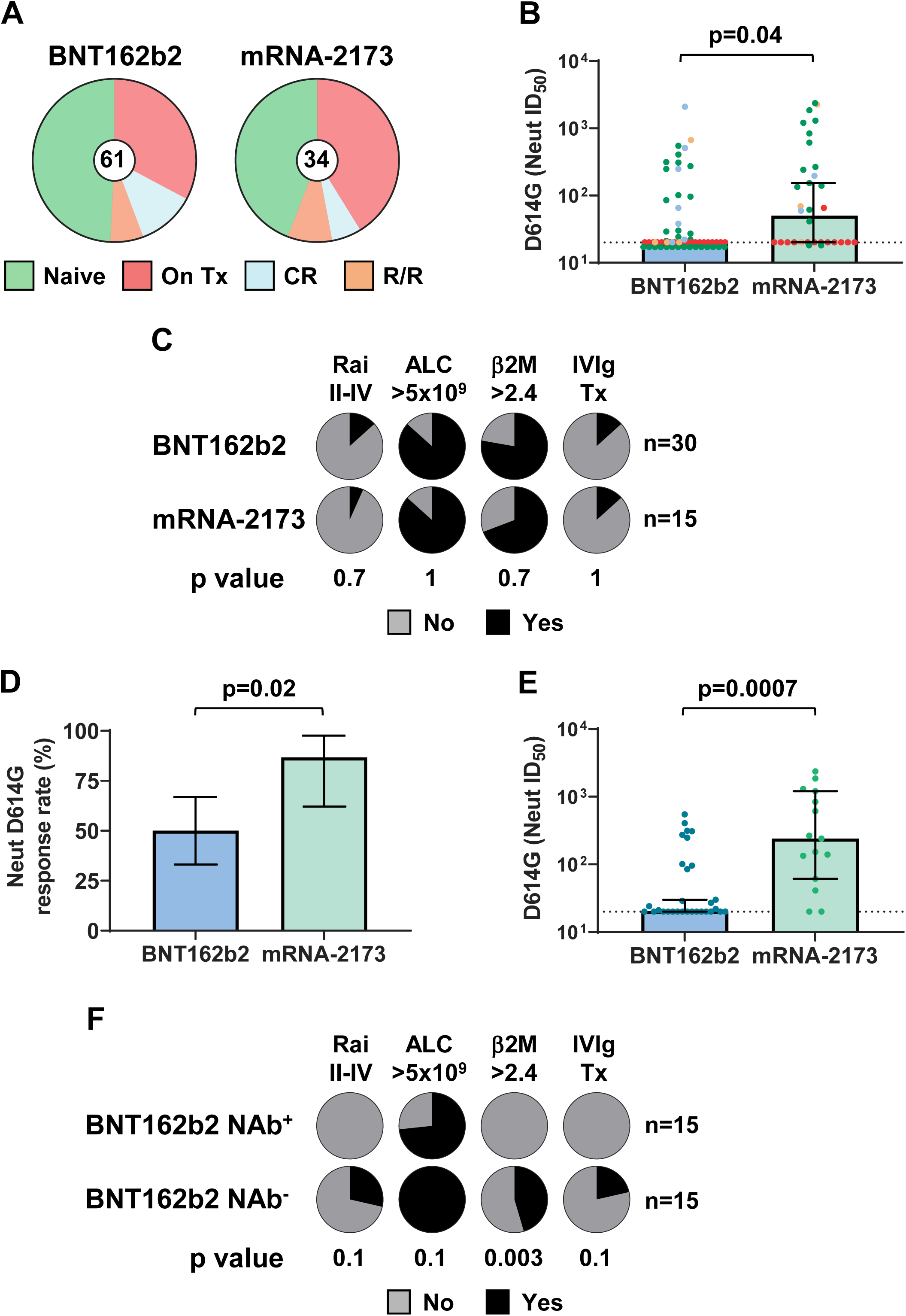
NAb titers are higher in treatment-naive mRNA-2173 CLL vaccinees, but impaired in patients with features of clinical progression. (A) BNT162b2 (n=61) and mRNA-2173 (n=34) CLL vaccinees stratified by treatment-naive (Naive), active treatment (On Tx), clinical remission (CR), or relapsed/refractory (R/R) disease status. (B) Comparison of ID_50_ neutralizing titers against the SARS-CoV-2 D614G variant for CLL patients classified by disease status (color coded as in A) and BNT162b2 or mRNA-2173 vaccine type. (C) Frequencies of four clinical features including Rai stage II-IV, absolute lymphocyte count (ALC; >5 x 10^9^/L), serum beta 2 microglobulin (β2M; > 2.4 mg/L), and IVIg prophylaxis therapy, among treatment-naive CLL patients by BNT162b2 or mRNA-2173 vaccine type. (D) Response rates for the generation of D614G NAbs by vaccine type in treatment-naive CLL patients. (E) Comparison of ID_50_ titers against D614G for treatment-naive CLL patients by vaccine type. (F) Frequencies of four clinical features between treatment-naive BNT162b2 vaccinees stratified by NAb^+^ (n=15) or NAb^-^ (n=15) serologic status. Bars indicate the mean (D) or median with 95% CI (B and E). P values were calculated with Fisher’s exact test (C-D, F) or the Mann-Whitney test (B and E).

That fact that there were comparable numbers of D614G NAb^+^ (n=15) and NAb^-^ (n=15) individuals who received the BNT162b2 vaccine provided an opportunity to examine reasons for these differences. The average time from the second vaccination to sample collection did not differ between the NAb^+^ and NAb^-^ groups (64.7 vs 67.3 days). However, comparisons of clinical parameters indicated a trend toward more advanced disease in NAb^-^ CLL vaccinees, with higher Rai stage, increased serum β2M levels, and IVIg requirements that were absent in NAb^+^ CLL vaccinees (Figure 4F). These disease characteristics also correlated with poor D614G NAb responses when the entire CLL cohort was analyzed (Table S4A). Among treatment-naïve BNT162b2 vaccinees, elevated serum β2M was associated with a 3.5-fold higher risk of failing to mount a D614G NAb response (95% CI, 1.8-7.2, p = 0.003). Due to limited sample numbers, Rai stages II-IV and IVIg therapy alone were not predictive, but when considered in combination, their presence predicted a 2.7-fold higher risk of failing to develop NAbs (95% CI, 1.4-4.7, p = 0.02). Similar results were obtained when NAb titers to the Delta variant were compared, with elevated serum β2M conferring a 2.6-fold higher risk (95% CI, 1.4-4.8, p = 0.02) and the combination of Rai stages II-IV and IVIg therapy conferring a 2.4-fold higher risk (95% CI, 1.5-4.3, p=0.004) of impaired NAb responses (Figure S3D). These findings indicate that CLL vaccinees who are unable to develop NAb responses possess features of advanced clinical disease.

### Treatment-naïve CLL vaccinees who are unable to mount NAb responses have lower CD4^+^ T cells

Interactions between CD4^+^ T and B cells are critical for germinal center reactions.^40, 41^ Because these lymphocyte subsets decline as a function of both age and CLL disease,^2, 42, 43^ we compared T cell frequencies in age-matched healthy controls (n=7) as well as treatment-naïve SARS-CoV-2 vaccinees who did (S^+^NAb^+^, n=11) versus did not (S^+^NAb^-^, n=9) mount a neutralizing antibody response. As observed for the overall CLL cohort (Figure S1), total CD3^+^ T cells were significantly reduced in S^+^NAb^+^ and S^+^NAb^-^ patients relative to controls (Figure 5A). However, S^+^NAb^-^ vaccinees had significantly lower numbers of total CD4^+^ T cells and a trend toward higher CD8^+^ frequencies, as reflected by lower CD4:CD8 ratios (Figures 5B-5D). Similar trends were evident for naïve and memory CD4^+^ and CD8^+^ subsets. Compared to controls, naïve CD4^+^ T cells were significantly lower in S^+^NAb^-^, but not S^+^NAb^+^ patients, while there was a coincident rise in effector memory CD8^+^ T cells in the former group (Figures 5E and 5F). These data indicate an association between lower naïve CD4^+^ T cell numbers and the inability of treatment-naive CLL patients to generate NAb responses.

**Figure 5.**
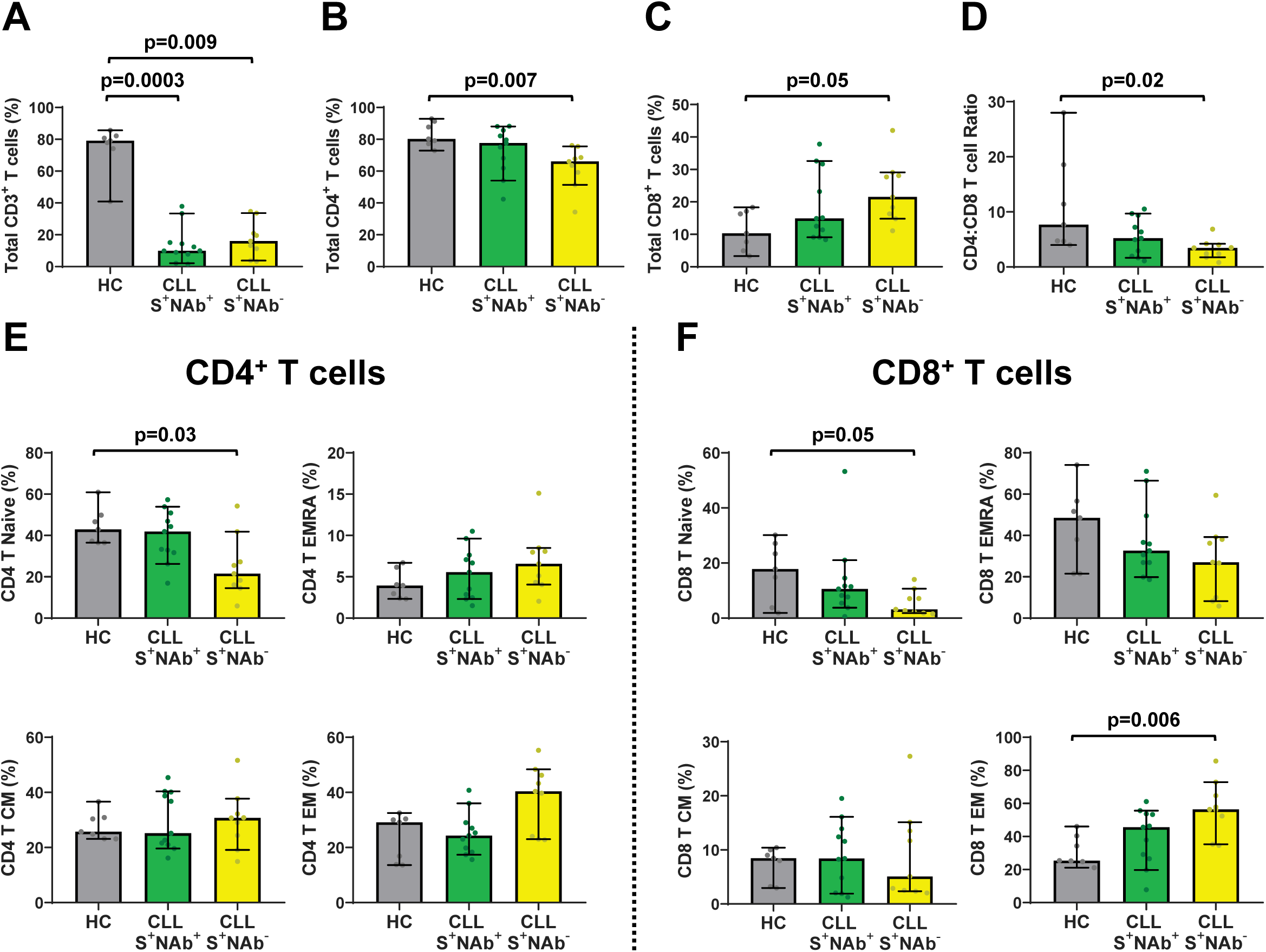
Progressive alterations in naive CD4^+^ and effector memory CD8^+^ T cells correlate with a loss of NAb responses in treatment-naive CLL vaccinees. (A-F) PBMCs from vaccinated healthy controls ≥ 65 years old (n=7) and treatment-naive CLL donors with the S^+^NAb^+^ (n=11) or S^+^NAb^-^ (n=9) serologic profiles were stained to enumerate the frequencies of total CD3^+^ (A), CD4^+^ (B), and CD8^+^ (C) T cells and CD4:CD8 ratios (D) as well as CD4^+^ (E) or CD8^+^ (F) naïve (N), central memory (CM), effector memory (EM) and effector memory CD45RA^+^ (EMRA) T cell subpopulations. Bars indicate the median with 95% CI. P values were determined by Dunn’s test of multiple comparisons following a Kruskal-Wallis test.

## DISCUSSION

CLL is a slowly advancing B cell lymphoproliferative disorder that ultimately impairs the ability to mount an effective immune response to new infections. Here, we compared the immunogenicity of BNT162b2 and mRNA-2173 SARS-CoV-2 vaccines in a clinically well-characterized cohort of CLL patients and discovered a previously unappreciated progressive loss of adaptive immune functions in treatment-naive CLL patients (Figure 6). By examining both binding and neutralizing antibody responses, we found a subset of vaccinees that was still able to mount *de novo* responses to SARS-CoV-2 antigens, albeit at titers lower than healthy controls (S^+^NAb^+^, light green). A second group of CLL vaccinees was unable to generate SARS-CoV-2 neutralizing antibodies (S^+^NAb^-^, yellow), but had spike binding antibodies which primarily reacted with epitopes in the SARS-CoV-2 S2 subunit. Although these could be vaccine-induced SARS-CoV-2 directed antibodies, it is more likely that they represent recall responses of pre-existing anti-HCoV antibodies that cross-react with conserved S2 epitopes. The latter possibility is reminiscent of the concept of antigenic imprinting or original antigenic sin,^44^ which refers to the preferential reactivation of cross-reactive memory B cells from an initial antigenic exposure, rather than the initiation of *de novo* responses when encountering a new related antigen. The fact that S^+^NAb^-^ vaccinees had more advanced disease with lower naïve CD4^+^ and higher CD8^+^ effector memory T cells is consistent with this interpretation. The third group of CLL vaccinees had no detectable SARS-CoV-2 antibodies (S^-^NAb^-^, light red), indicating an inability to mount *de novo* as well as recall responses. Most of these individuals required IVIg prophylaxis, demonstrating they were the most immune compromised. Thus, SARS-CoV-2 vaccination exposed a progressive loss of immune functions in CLL patients, with pre-existing memory being preserved longer than the capacity to respond to new antigens (Figure 6).

**Figure 6.**
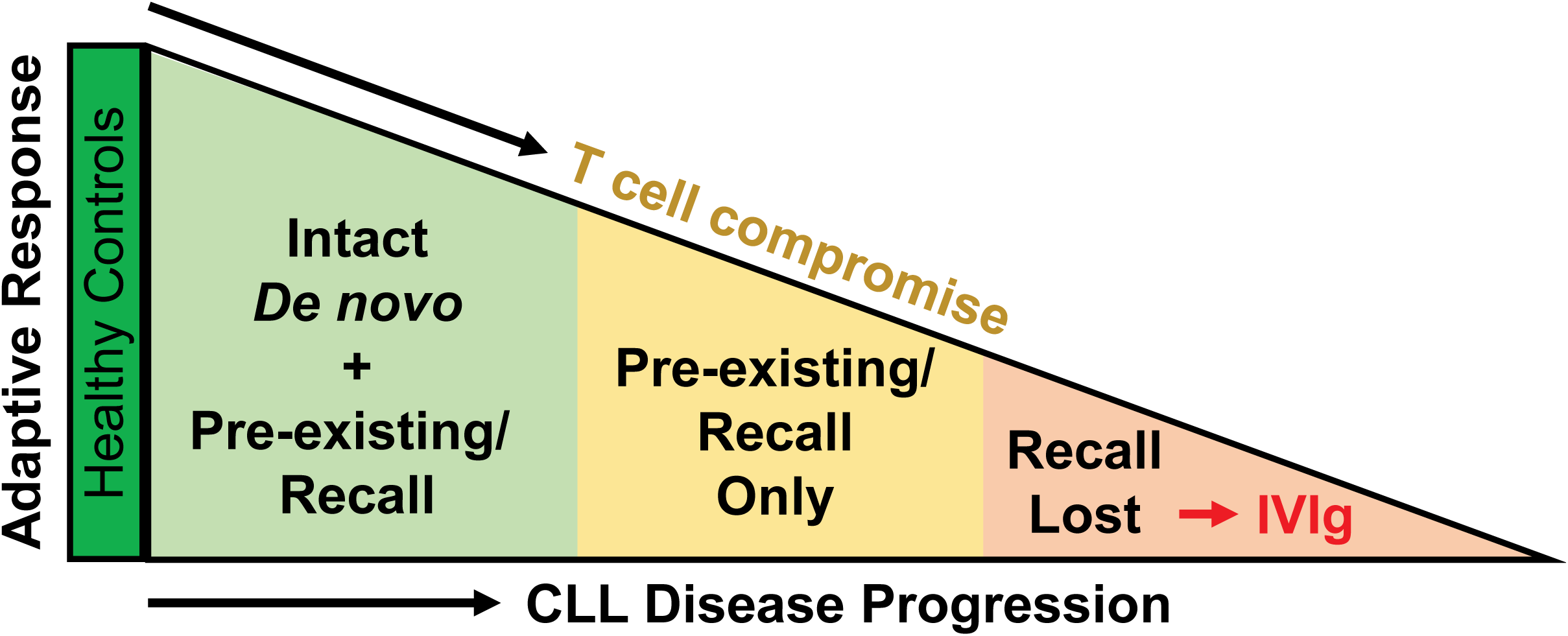
Progressive loss of adaptive immunity in treatment-naive CLL patients. Humoral and cellular immune responses following SARS-CoV-2 mRNA vaccination uncovered three distinct serologic profiles in CLL patients compared to healthy controls that reflect a graded decline in adaptive immune function.

Although we cannot exclude that S^+^NAb^-^ CLL patients mounted some *de novo* responses, the absence of detectable neutralizing antibodies indicates that these individuals lacked key immune elements required for the induction of germinal center B cell responses and antibody affinity maturation. This disparate humoral response was accompanied by diminished frequencies and altered functions of T cells that were more biased towards a CD8^+^ response. Nonetheless, S^+^NAb^-^ CLL vaccinees maintained HCoV-specific antibodies at levels comparable to healthy controls. Thus, SARS-CoV-2 vaccination in the context of a partially compromised immune system may favor reactivation of pre-existing memory over the stimulation of naïve B cells, similar to what has been observed for responses to influenza following vaccination in the elderly.^43^ Given the essential contributions of naïve B and CD4^+^ T cells to *de novo* responses, their decline over the CLL disease course is expected to worsen the capacity for engaging neoantigens. Germinal center-based functions would be increasingly diminished and the potential for generating new responses would eventually be lost. Our study thus suggests that vaccination against SARS-CoV-2 or other neoantigens could be used as a tool to assess the status of this decline and to gain greater insight into the underlying mechanisms contributing to the immune impairment.

Recent studies have shown that CLL patients benefit from adjuvanted zoster vaccines, including individuals on BTK inhibition, but that hepatitis B immunization elicited poor or no responses in both treatment-naive and BTK inhibitor treated patients.^8^ The effectiveness of pneumococcal and influenza immunization is also low in treatment-naive CLL patients, but has been observed to improve with higher dosing, adjuvant-conjugation, and earlier administration following diagnosis.^6, 7, 45^ These results suggest that CLL patients with a reduced ability to mount *de novo* responses may benefit from more rationally designed vaccine regimens. Indeed, about a quarter of CLL patients who failed to respond to two SARS-CoV-2 mRNA immunizations, subsequently seroconverted following a third immunization.^46, 47^ Thus, CLL patients should be considered candidates for alternative vaccination strategies. For example, the high-dose flu vaccine, which is tailored for the elderly, may elicit stronger and broader responses in CLL patients with partially impaired adaptive immunity. Similarly, a COVID vaccine that includes more than one variant may improve *de novo* responses. Clinical trials that formally test these possibilities should be of high priority.

For treatment-naive patients who lack humoral responses (S^-^NAb^-^) or those on B cell targeted therapies that inhibit seroconversion or NAb generation, passive immunotherapies and antivirals will continue to be important. More than 25% of CLL patients require IVIg^9^ and recent studies confirm the presence of NAbs in US preparations.^48^ Prophylactic administration of recombinant NAbs have also proved beneficial in the immunocompromised.^49^ However, it is difficult for these treatments to keep up with the pace of viral diversification and resistance to antivirals will likely also occur.^50^ Given the benefits of polyclonal convalescent plasma,^22^ a cocktail of prophylactic recombinant NAbs engineered for breadth and durability will thus be a critical advance for this patient population.

A key finding in our study was the demonstration of superior neutralizing antibody responses in CLL patients who received the mRNA-2173 vaccine. This observation is consistent with previous findings of improved seroconversion rates and T cell activity in COVID vaccine studies of other hematologic malignancies, including CLL,^25, 47, 51^ as well as immunocompromised patients with non-malignant conditions such as solid organ transplant or rheumatologic disorders.^52^ The higher neutralizing response rates and titers in mRNA-2173 vaccinees may be due to the mRNA dose, which is ∼3.3-fold higher compared to BNT162b2.^53, 54^ Indeed, in the elderly, a 100 µg mRNA-1273 dose elicited higher binding and neutralizing antibody titers than a 25 µg dose.^55^ However, the two vaccines also differ in other properties, including their formulation.^56^ Regardless of the reasons, the fact that the mRNA-2173 vaccine elicited higher NAb titers in a larger fraction of CLL patients suggests that this vaccine may confer greater protection from SARS-CoV-2 in this vulnerable population.

In summary, our study of SARS-CoV-2 vaccine-induced humoral and cellular responses in CLL patients provides a more nuanced picture of their inherent immune dysfunction, with pre-existing immunity being preserved longer than the capacity to mount *de novo* responses. Moreover, higher NAb titers and response rates identify mRNA-2173 as a potentially superior vaccine in CLL patients. Future studies should consider the utility of vaccination to assess the extent of disease-induced immune dysfunction in CLL patients and to gain greater insight into the underlying mechanisms.

## Data Availability

All relevant data are within the manuscript and its Supporting Information files.

## ACKNOWLEDGEMENTS

We thank the donors and patients who contributed samples as well as the UAB Hematology/Oncology staff who supported this study. These studies were funded in part by the UAB School of Medicine Dean’s office COVID-19 research initiative, the UAB Cancer Immunobiology Program, and grants from the NIH (UM1 AI069452, R01 AI150590, and U01 CA260462).

## AUTHOR CONTRIBUTIONS

Conceptualization, A.B., P.A.G., B.H.H., and R.S.D.; Methodology, K.Q., K.H., W.L., A.B., T.J.K., A.C., V.M.C-P., B.M.L., and R.S.D.; Validation, K.Q., K.H., W.L., R.S., L.A.H., A.K.O., and S.S.M.; Formal Analysis, K.Q., K.H., S.S.M., and R.S.D.; Investigation, K.Q., K.H., S.S.M., W.L., R.S., A.K.O., and L.A.H.; Resources, R.L., S.S., E.R.F., J.R.L., M.N., A.M., F.J.O., R.B.P., T.J.K., B.M.L., P.A.G., B.H.H., and R.S.D.; Writing, B.H.H. and R.S.D.

## METHODS AND MATERIALS

### Human samples

Blood samples were collected following institutional review board (IRB) approval by the University of Alabama at Birmingham (IRB #130821005 and 160125005) and written informed consent was obtained from all participants who were adults over age 18 and recruited between January and August 2021. Healthy controls (n=30) and CLL patients (n=95), who were diagnosed according to the International Workshop on Chronic Lymphocytic Leukemia guidelines,^3^ were vaccinated in the community with the first and second doses of the Pfizer/BioNTech (BNT162b2) or Moderna (mRNA-1273) COVID-19 vaccines. Human peripheral blood mononuclear cells (PBMCs) and plasma samples were processed as previously described.^22, 57^ Details of the vaccinated CLL and healthy adult cohorts are listed in Tables 1 and S1. Samples were anonymized and coded so that any patient/participant/sample identifiers included were not known to anyone (e.g., hospital staff, patients or participants themselves) outside the research group so cannot be used to identify individuals.

To confirm that healthy donors and CLL patients had not been recently infected with SARS-CoV-2, blood samples were analyzed for the presence of nucleocapsid IgG antibodies in a clinical diagnostic laboratory (Abbott).^58^

Clinical characteristics of CLL patients were extracted by retrospective analysis of the electronic medical record and included demographics, disease and therapeutic history, Rai stage, laboratory data of the complete blood count, serum beta 2 microglobulin (β2M) and immunoglobulin levels, immunoglobulin heavy chain variable gene (*IGHV*) mutation status (delineated by germline identity of 98%), CD38 expression (≥20%), and cytogenetics analysis by fluorescence *in situ* hybridization (FISH).^59^

### SARS-CoV-2 S-protein and subunit ELISA

Plasma IgG binding antibodies to the entire SARS-CoV-2 spike (S) protein as well as receptor binding domain (RBD), and S1 and S2 proteins, were detected by enzyme-linked immunosorbent assay (ELISA) using recombinantly expressed (Wuhan-Hu-1 S-protein and RBD) and purchased (Wuhan-Hu-1 S1 and S2 proteins, ACROBiosystems) proteins as previously described.^27, 60, 61^ Briefly, Costar high binding flat-bottom 96-well plates were coated with 300 ng per well of a pre-fusion stabilized (S-2P) S protein (residues 1–1138) (plasmid kindly provided by Philip Brouwer and Rogier W. Sanders, Department of Medical Microbiology, University of Amsterdam, Amsterdam, The Netherlands) or 400 ng of recombinantly expressed RBD (residues 419–541), S1 (residues 16-685) or S2 protein (residues 686-1213 with F817P, A892P, A899P, A942P, K986P and V987P substitutions for stabilization) in PBS overnight at 4°C and then incubated with blocking buffer (5% non-fat milk powder in PBS + 0.05% Tween 20) for 1 h at 37°C. Plasma samples were heat-inactivated at 56°C for 1 hour, 5-fold serially diluted in blocking buffer and then added to the plates for 1 h at 37°C. After five washes with PBS-T (PBS + 0.1% Tween 20), plates were incubated for 1 h at 37°C with horseradish peroxidase (HRP)-conjugated goat-anti-human IgG detection antibodies diluted 1:5,000 in blocking buffer. After five additional washes, 3, 3′, 5, 5′-tetramethylbenzidine (TMB) substrate was added for color development for 10 min before the reaction was stopped with an equal volume of 1N H2SO4. Absorbance was read at 450 nm using a Synergy 4 spectrophotometer. The average OD450 value from three background control wells (no plasma) was subtracted from the protein coated wells. In addition, the average OD450 value (plus two standard deviations) of 28 pre-pandemic sera was subtracted from each plasma dilution. Midpoint (EC_50_) and endpoint titers were determined as described.^22, 27^ Briefly, midpoint (EC_50_) titers were calculated by a nonlinear-regression fit of a 4-parameter sigmoid function to the corrected OD450 values and the logarithmic dilution factors (the lower plateau was set to 0; GraphPad Prism software). End-point titers were read from the fitted curve at a corrected OD450 cutoff of 0.1.

### SARS-CoV-2 pseudovirus neutralization assay

Plasma samples of vaccinees were tested for neutralizing responses against the SARS-CoV-2 variants D614G^28^ and B.1.617.2 (also termed Delta with mutations T19R, G142D, ΔE156, ΔF157, R158G, L452R, T478K, D614G, P681R, and D950N compared to Wuhan-Hu-1),^29, 30^ using an HIV-1 based pseudovirus assay as previously described.^31^ Briefly, pseudovirus stocks were generated by co-transfecting spike expression plasmids (encoding proteins with a 19 amino acid cytoplasmic tail deletion) with an HIV-1 nanoluciferase encoding reporter backbone in HEK293T cells. Pseudovirus stocks were tittered to identify the appropriate infectious dose, incubated with five-fold serial dilutions of vaccinee plasma and then used to infect 1.5 x 10^4^ 293T clone 13 cells expressing ACE2. Two days post-infection, cells were washed with PBS, lysed, and nanoluciferase activity was determined using a Nano-Glo® Luciferase Assay System. Luciferase activity in wells with virus and no plasma were set to 100%, and the dilution of plasma at which luminescence was reduced to 50% (Inhibitory Dose 50; ID_50_) was calculated as an average of two technical replicates. Each vaccinee plasma was analyzed under anonymized code on at least two occasions, with the geometric mean of all measurements reported. Values below a titer of 1:20 were treated as 20 when averaging.

### ACE2/RBD binding inhibition assay

Plasma samples were analyzed with an ACE2/receptor binding domain (RBD) binding inhibition assay as previously described.^22, 62^ High-binding 96-well plates (Corning #3690) were coated with 50 µl per well of recombinant RBD (Wuhan-Hu-1, RayBiotech) diluted at 1 µg/ml in PBS at 4°C overnight. The following day, plates were washed 3 times with PBS + 0.1% Tween 20 (PBST), and wells were blocked with 100 µl per well of 3% non-fat dry milk in PBST by incubation at room temperature for 1 h. After washing the blocked wells 3 times with PBST, either 50 µl of plasma serially diluted in 1% non-fat dry milk in PBST, or 1% non-fat dry milk in PBST alone as a no inhibition control, was added to wells, and incubated at room temperature for 2 h. Heat-inactivated plasma samples (56°C for 30 min) were initially diluted at 1:25, then serially diluted 2-fold for the assay to 1:400. After incubation, plates were washed 3 times with PBST, then 50 µl of recombinant human ACE2 (RayBiotech) diluted at 0.1 µg/ml in PBST was added to the wells. Plates were incubated at room temperature for 1 h, washed 4 times with PBST, and 50 µl of biotinylated goat anti-human ACE2 (R&D) diluted at 0.1 µg/ml in PBST was added to the wells. Plates were incubated at room temperature for 1 h, washed 4 times with PBST, and then 50 µl of HRP-conjugated streptavidin (Southern Biotech) (1:2,000 in PBST) was added to the wells, and incubated at room temperature for 30 min. Plates were washed 5 times with PBST, developed with 50 µl per well of 3,3′,5,5′-tetramethylbenzidine TMB substrate (Biolegend) at room temperature for 8 min, and the reaction was stopped by addition of 50 µl of 1N H2SO4. The OD was measured at 450 nm with a SPECTROstar omega (BMG Labtech) microplate reader. ACE2 binding was expressed as a percentage of OD values relative to the OD450 value of a no inhibition control. Binding values of <90% at a 1:25 dilution of plasma were used to calculate inhibitory activity. The upper limit of the assay was 100%.

### Human Coronavirus (HCoV) spike protein ELISA

High-binding 96-well plates (Corning #3690) were coated at 4°C overnight with 50 µl per well of the following recombinant Spike (S1+S2) proteins: SARS-CoV, MERS-CoV, HCoV-HKU1, HCoV-OC43, HCoV-NL63, or HCoV-229E (all from Sino Biological) diluted at 2 µg/ml in PBS. The following day, plates were washed 3 times with 0.1% Tween 20 in PBS (PBST). A blocking solution of 100 µl per well of 3% non-fat dry milk in PBST was added and incubated at room temperature for 1 h before washing plates 3 times with PBST. Plasma samples were heat-inactivated at 56°C for 30 min and initially diluted at 1:20, then serially diluted 5-fold in 1% non-fat dry milk in PBST before adding 50 µl per well, and incubating at room temperature for 2 h. Plates were then washed 4 times with PBST, and 50 µl per well of horseradish peroxidase (HRP) conjugated goat anti-human IgG (Southern Biotech #2045-05) diluted at 1:6,000 in PBST was added to the wells and incubated at room temperature for 1 h to measure Spike protein IgG Ab responses. Plates were washed 5 times with PBST, developed with 50 µl per well of HRP substrate 3, 3’, 5, 5’ tetramethyl benzidine (TMB, Biolegend) at room temperature for 10 min.

The reaction was stopped by addition of 50 µl per well of 1N H2SO4. The optical density (OD) was measured at 450 nm with an Epoch microplate spectrophotometer (BioTek). OD450 values of blank wells, which were Spike protein-coated wells without plasma, were subtracted from OD450 values of sample wells, and EC_50_ values were determined by a nonlinear-regression fit of a 4-parameter sigmoid function with the corrected OD450 values and the dilution factors.

### Flow cytometry-based T cell immunophenotyping, AIM, and ICS analyses

For immunophenotyping, cryopreserved PBMCs were thawed and washed with FACS wash buffer (2% FBS in PBS). Cells were then stained with CCR7-PerCP5.5, incubated at 37°C for 20 min, and then stained with the following antibodies: CD3-Alexa780, CD4-BV711, CD8-FITC, CD45RA-BV510, CD19-BUV563, and LIVE/DEAD-UV. After incubation at 4°C for 30 min, cells were washed twice with FACS wash buffer (2% FBS in PBS) and fixed in a 4% formalin solution. Events were collected on a BD FACSymphony A3 instrument within 24 h and analyzed using FlowJo software (v10).

For activation-induced marker staining (AIM), antigen-specific T cells were measured as previously described.^57^ PBMCs were stimulated with SARS-CoV-2 N and S protein peptide pools (BEI Resources) at an individual peptide concentration of 1 µg/ml in the presence of co-stimulatory anti-CD28 and anti-CD49d antibodies (BD Pharmingen). Cell aliquots from each sample were stimulated with equal amounts of dimethyl sulfoxide (DMSO) as a negative control and staphylococcal enterotoxin B (SEB) as a positive control. After incubation at 37°C for 18 h, cells were washed and stained with the following antibodies: CD4-BV711, CD3-Alexa780, CD8-FITC, CD19-BUV563, OX40-PECy7, PDL1-PE, CXCR5-BV421, PD1-BV785, CD137-BV650, CD69-BUV737, and LIVE/DEAD-UV. Cells were then washed and fixed in 4% formalin. Events were collected on a BD FACSymphony A3 instrument and analyzed using FlowJo software (v10).

Intra-cellular staining (ICS) experiments were performed in parallel with the AIM analysis as previously described.^57^ CD107a-FITC was added with the co-stimulatory antibody mix. Cells were incubated for a total of 12 h in total. Staining was conducted in three steps: 1) Surface marker staining for 30 min at 4°C with LIVE/DEAD-UV, CD3-Alexa780, CD4-BV711, CD8-V500, CD14-PercpCy5.5, and CD19-BUV563; 2) Permeabilization with CytoFix/CytoPerm solution (BD Biosciences) for 20 min at 4°C; and 3) ICS for 30 min at 4°C with IFNγ-Alexa700, TNFα-PECy7, IL2-APC, GranzymeB-V450, and Perforin-PE. Finally, cells were washed twice and fixed in 4% formalin. Events were collected on a BD FACSymphony A3 instrument and analyzed using FlowJo software (v10). For both AIM and ICS, positive responses were determined by comparison to an unstimulated control with a threshold above at least three times and higher statistical significance by calculation of Chi-square analysis with Yates’ correction (p value < 0.05). Combinatorial polyfunctionality analysis (COMPASS) of antigen-specific T-cells was calculated as previously described.^39^

### Statistical analysis

Data were analyzed in R v4.0.5 and GraphPad Prism version 9.0 software. Statistical details of the experiments are provided in the respective figure legends and tables. Associations between serological or cellular responses with dichotomous clinical data were analyzed using Fisher’s exact test and with continuous clinical variables using the Mann-Whitney test. P values were adjusted for multiple comparisons of serologic, cellular, and clinical data using the Benjamini– Hochberg procedure for false discovery rate. For comparisons of more than two categories, P values were determined by Dunn’s test of multiple comparisons following a Kruskal-Wallis test. Firth logistic regression was used to examine the association of serologic or cellular responses with clinical variables. GraphPad Prism version 9.0 software was used to plot these analyses. Significance was determined as p-value < 0.05 unless otherwise stated.

## FIGURE LEGENDS

**Figure S1.**
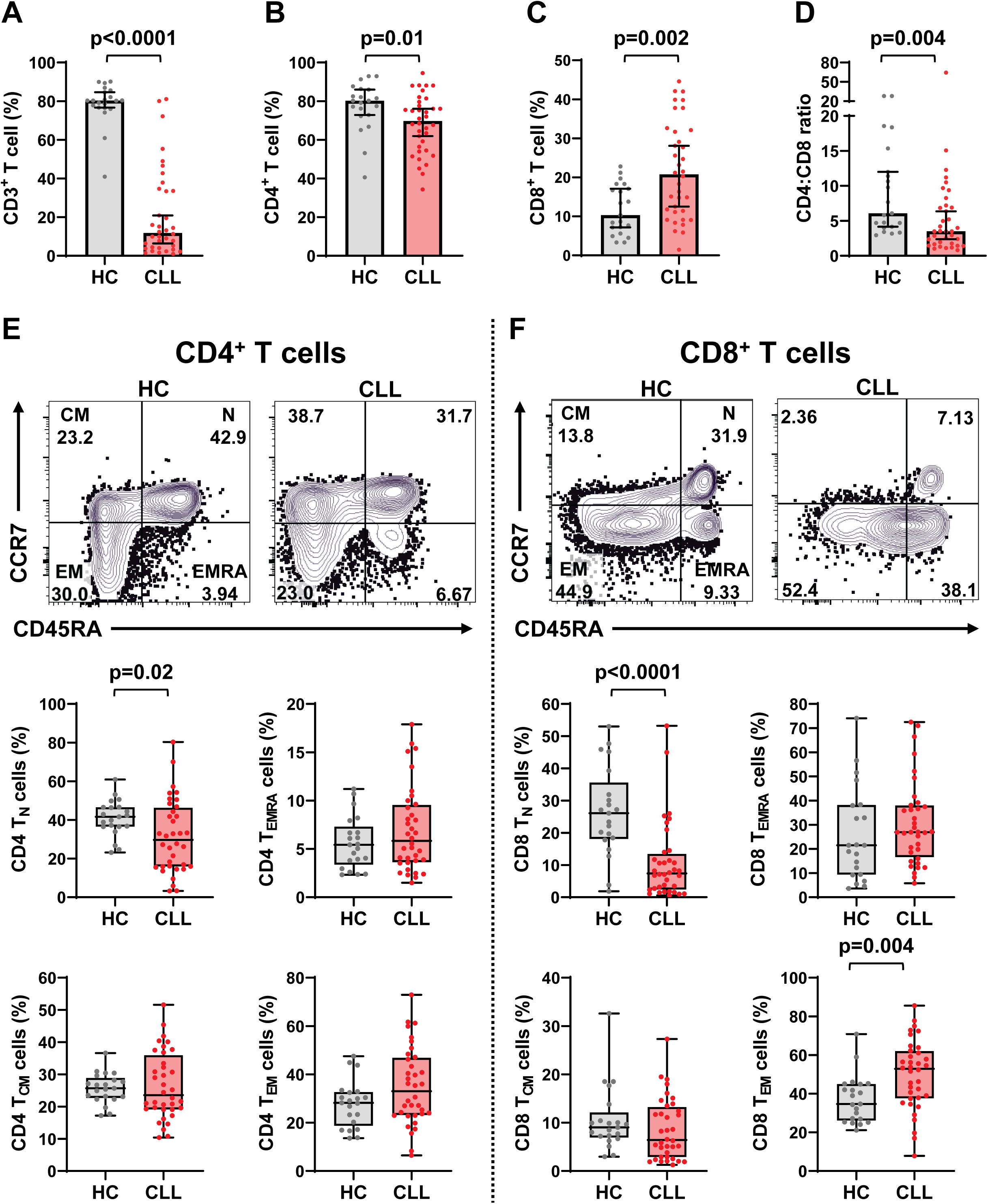
CLL patients have altered total CD4^+^ and CD8^+^ T cell and subpopulation frequencies, Related to Figures 2 & 5. Immunophenotyping of PBMCs from vaccinated HC (n=21) and CLL (n=36) donors. (A-D) Quantitative comparisons of total CD3^+^ (A), CD4^+^ (B), and CD8^+^ (C) T cell frequencies and CD4:CD8 ratios (D). (E-F) Representative flow cytometry plots and quantitative comparisons of naïve (N), central memory (CM), effector memory (EM) and effector memory CD45RA^+^ (EMRA) CD4^+^ (E) and CD8^+^ (F) subpopulation frequencies defined by the CCR7 and CD45RA surface markers in HC and CLL donors. Bars indicate the median with 95% CI. P values were determined by the Mann-Whitney test.

**Figure S2.**
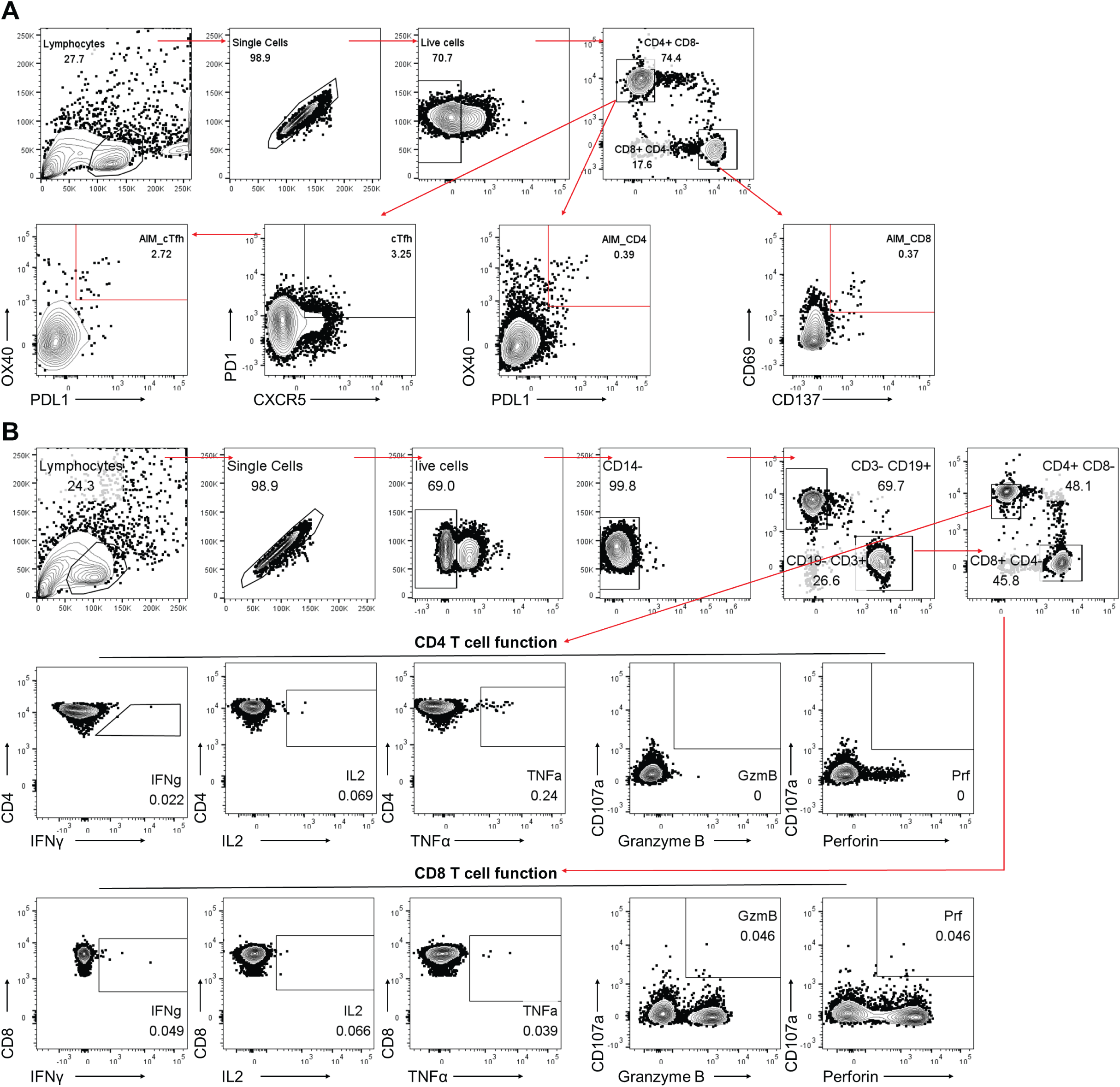
Flow cytometry gating strategies for measuring T cell responses from a CLL donor after S peptide pool stimulation, Related to Figure 2. (A) Gating strategy to examine activation-induced markers (AIM) by CD4+, cTfh, and cos+ T cells. (B) Gating strategy to examine CD4+ and COS+ T cell effector function by intra-cellular staining (ICS).

**Figure S3.**
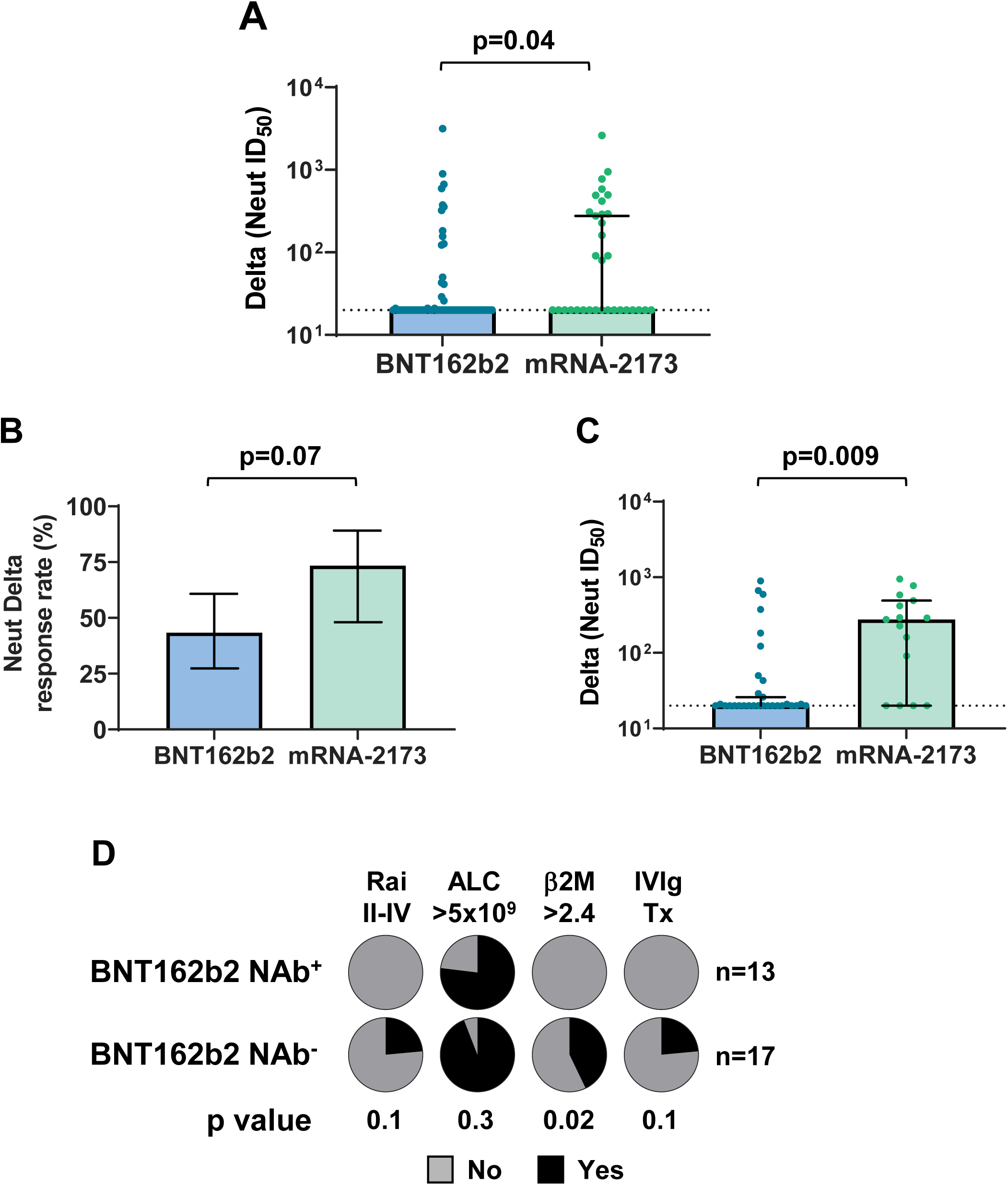
NAb titers against the Delta variant are higher in total and treatment-naïve mRNA-2173 CLL vaccinees, but lower in patients with clinical progression, Related to Figure 4. (A) Comparison of ID_50_ neutralizing titers against the SARS-CoV-2 Delta variant for CLL patients classified by BNT162b2 (n=60) or mRNA-2173 (n=33) vaccine type. (B) Response rates for the generation of Delta NAbs by vaccine type in treatment-naive CLL patients. (C) Comparison of ID_50_ titers against Delta for treatment-naive CLL patients by BNT162b2 (n=30) or mRNA-2173 (n=15) vaccine type. (D) Frequencies of four clinical features between treatment-naive BNT162b2 vaccinees stratified by Delta NAb^+^ (n=13) or NAb^-^ (n=17) serologic status. Bars indicate the median with 95% CI (A and C) or mean (B). P values were calculated with the Mann-Whitney test (A and C) or Fisher’s exact test (B and D).

**Table S1.**
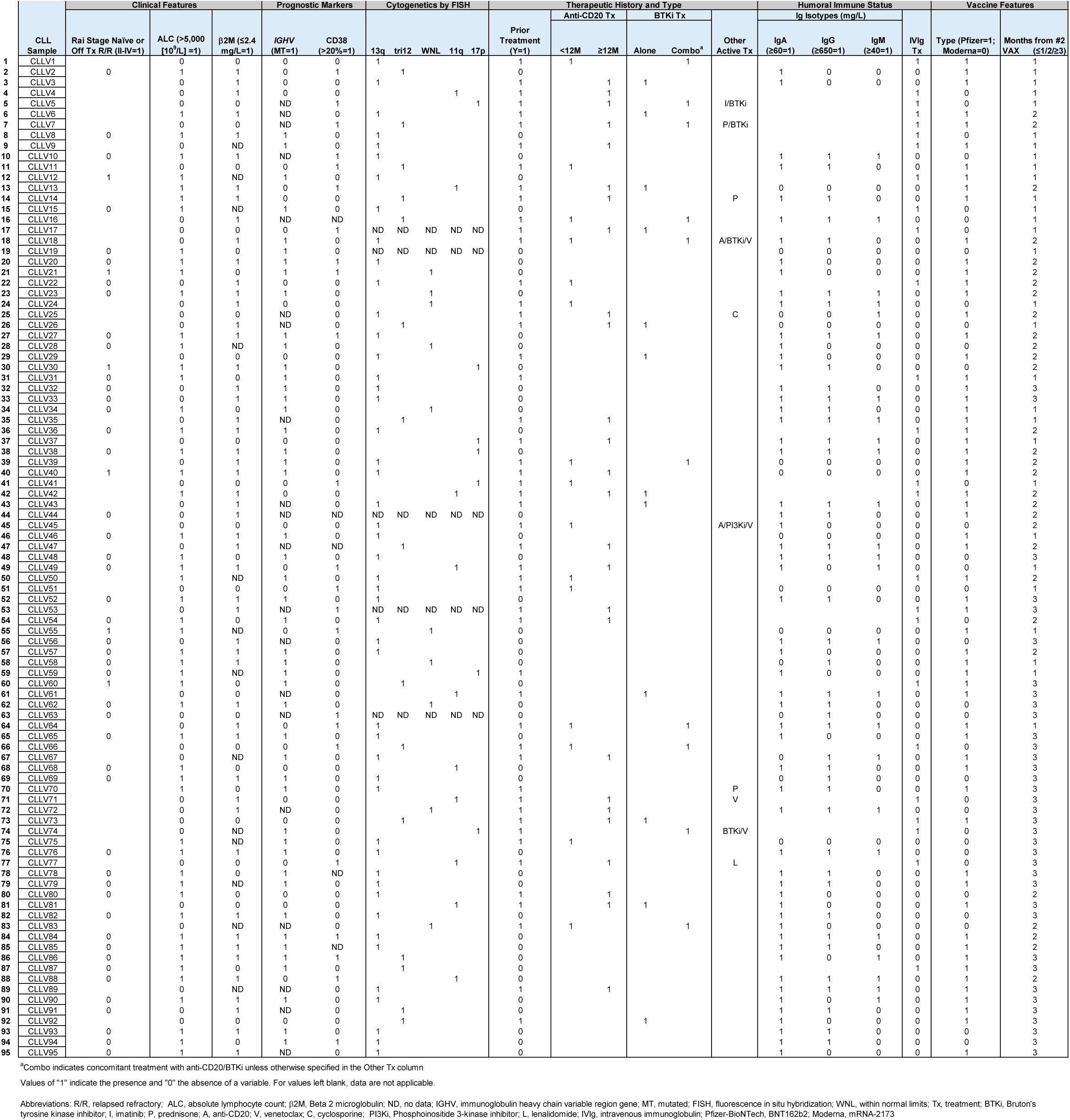
Disease characteristics and features of SARS-CoV-2 vaccinated CLL subjects, Related to Figures 1 & 4 and Tables 1 & S4.

**Table S2.**
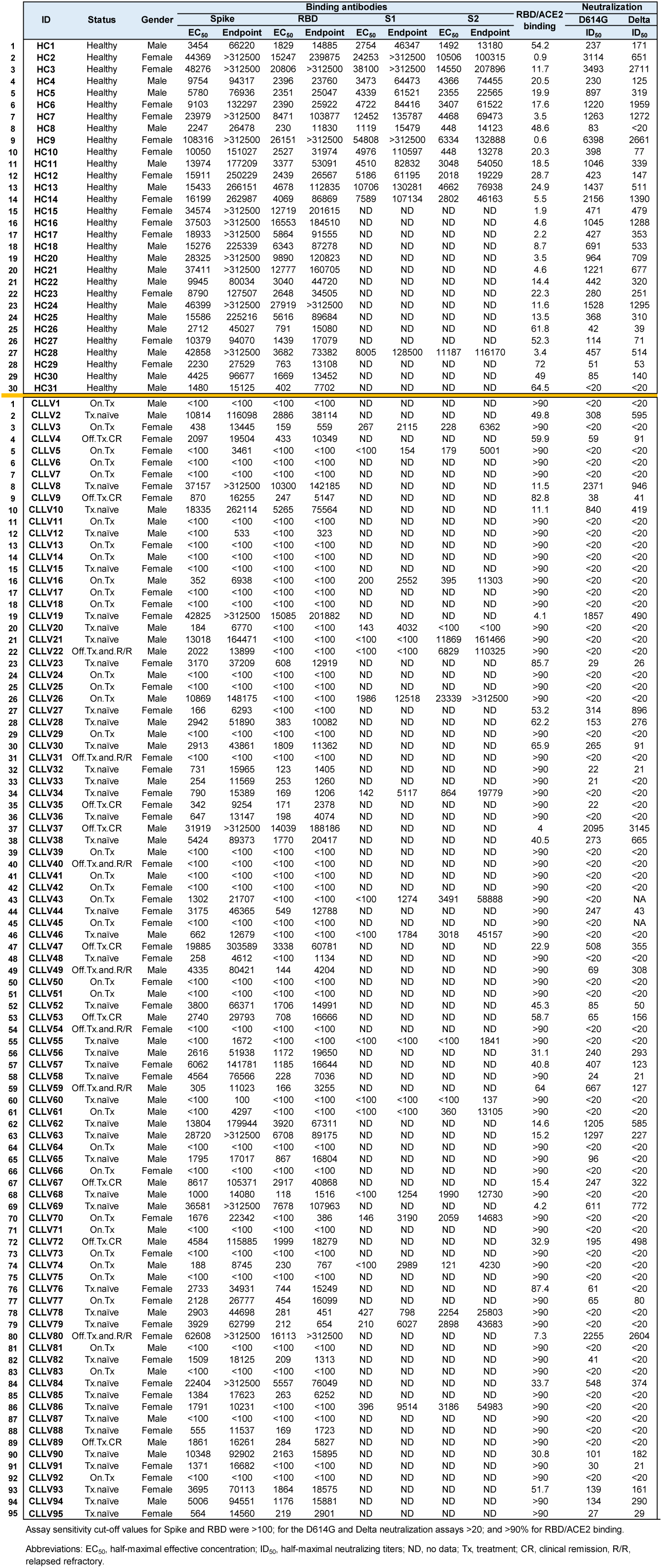
Binding and neutralizing antibody titers in the plasma of SARS-CoV-2 vaccinated CLL patients and healthy controls, Related to Figures 1, 3 & 4.

**Table S3.**
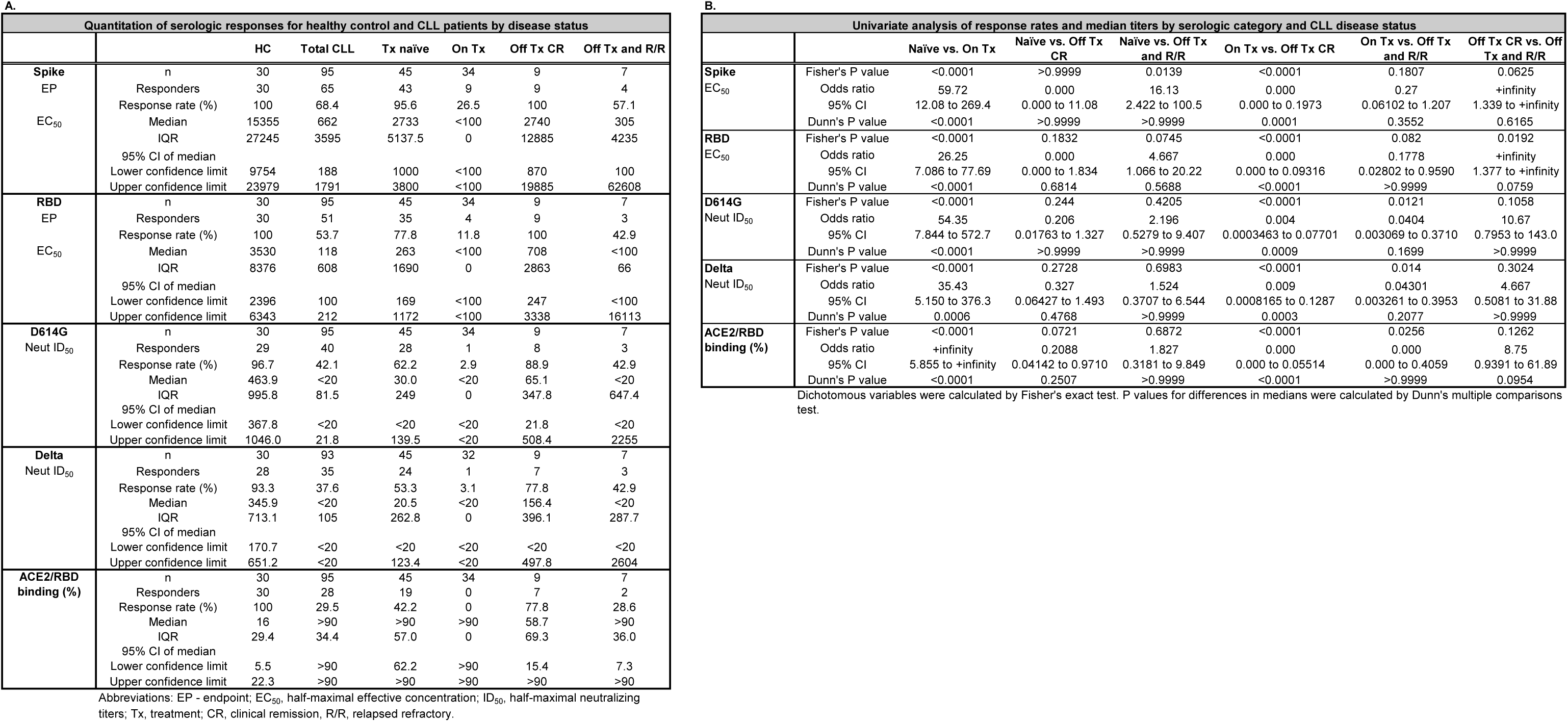
Serologic responses and univariate analyses for SARS-CoV-2 vaccinated healthy controls and CLL patients by disease status, Related to Figure 1.

**Table S4.** Clinical determinants of serologic and neutralizing responses in CLL vaccinees, Related to Figures 1 & 4.

**A. Categorical univariate analysis**
| Clinical Variable | Spike (EP) | RBD (EP) | D614G (Neut ID<sub>50</sub>) | Delta (Neut ID<sub>50</sub>) | ACE2/RBD Binding |
| --- | --- | --- | --- | --- | --- |
| Age ≥ 65 yo | NS | NS | NS | NS | NS |
| Gender | NS | NS | NS | NS | NS |
| Disease status | < 0.0001 | < 0.0001 | < 0.0001 | < 0.0001 | < 0.0001 |
| Rai stage II-IV (no) | NS | 0.038 | 0.034 | NS | NS |
| IGHV mutation status (MT) | 0.031 | 0.013 | NS | NS | NS |
| CD38 >20% | NS | NS | NS | NS | NS |
| FISH cytogenetics | NS | NS | NS | NS | NS |
| β2-microglobulin ≤ 2.4 | 0.0015 | 0.023 | < 0.0001 | 0.0094 | 0.013 |
| Prior therapy (no) | < 0.0001 | < 0.0001 | < 0.0001 | 0.0030 | 0.013 |
| Anti CD20 ≥ 12 months | 0.0082 | 0.0011 | 0.0030 | 0.0075 | 0.018 |
| BTK inhibitor therapy | NS | NS | NS | NS | NS |
| ALC < 5,000, (10<sup>9</sup>/L) | 0.047 | 0.014 | NS | NS | NS |
| IVIg prophylaxis (no) | 0.0011 | 0.019 | 0.010 | 0.052 | NS |
| IgG ≥ 650 mg/dL | 0.010 | 0.0027 | NS | NS | NS |
| IgM ≥ 40 mg/dL | NS | NS | NS | NS | NS |
| IgA ≥ 60 mg/dL | NS | 0.035 | NS | NS | NS |
| Pfizer or Moderna vaccine | NS | NS | NS | NS | NS |
| Months from vaccination | NS | 0.046 | NS | NS | NS |

**B. Multivariate analysis**
| Clinical Variable | Spike (EP) |  |  | D614G (Neut ID<sub>50</sub>) |  |  |
| --- | --- | --- | --- | --- | --- | --- |
|  | P value | OR | 95% CI | P value | OR | 95% CI |
| Age ≥ 65 yo | 0.26 | 0.42 | 0.079-1.9 | 0.87 | 1.1 | 0.29-4.5 |
| Male sex | 0.65 | 1.4 | 0.36-5.6 | 0.69 | 1.3 | 0.37-4.4 |
| Rai stage II-IV | 0.63 | 1.8 | 0.14-18 | 0.24 | 3.5 | 0.46-56 |
| Unmutated IGHV | 0.94 | 0.93 | 0.11-6 | 0.83 | 1.2 | 0.28-5.1 |
| On therapy | 0.0028 | 62 | 3.6-3500 | 0.041 | 40 | 1.2-2500 |
| Off therapy and R/R | 0.038 | 7.7 | 1.1-64 | 0.6 | 1.7 | 0.25-13 |
| Off therapy CR | 0.54 | 3.7 | 0.018-250 | 0.81 | 0.71 | 0.034-17 |
| ALC > 5,000, (10<sup>9</sup>/L) | 0.3 | 3.8 | 0.37-130 | 0.13 | 5.2 | 0.65-110 |
| IVIg prophylaxis therapy | 0.018 | 5.2 | 1.3-29 | 0.052 | 4.4 | 0.99-25 |
| Pfizer vaccination | 0.59 | 1.4 | 0.39-5.9 | 0.0056 | 5.8 | 1.6-27 |
| 2 months from vaccination | 0.3 | 2.3 | 0.49-12 | 0.9 | 1.1 | 0.24-5 |
| >3 months from vaccination | 0.51 | 0.59 | 0.11-2.8 | 0.91 | 1.1 | 0.25-5 |

Firth logistic regression was used to examine the association of serological responses with clinical variables.
Abbreviations: R/R, relapsed refractory; CR, clinical remission.

**Table S5.**
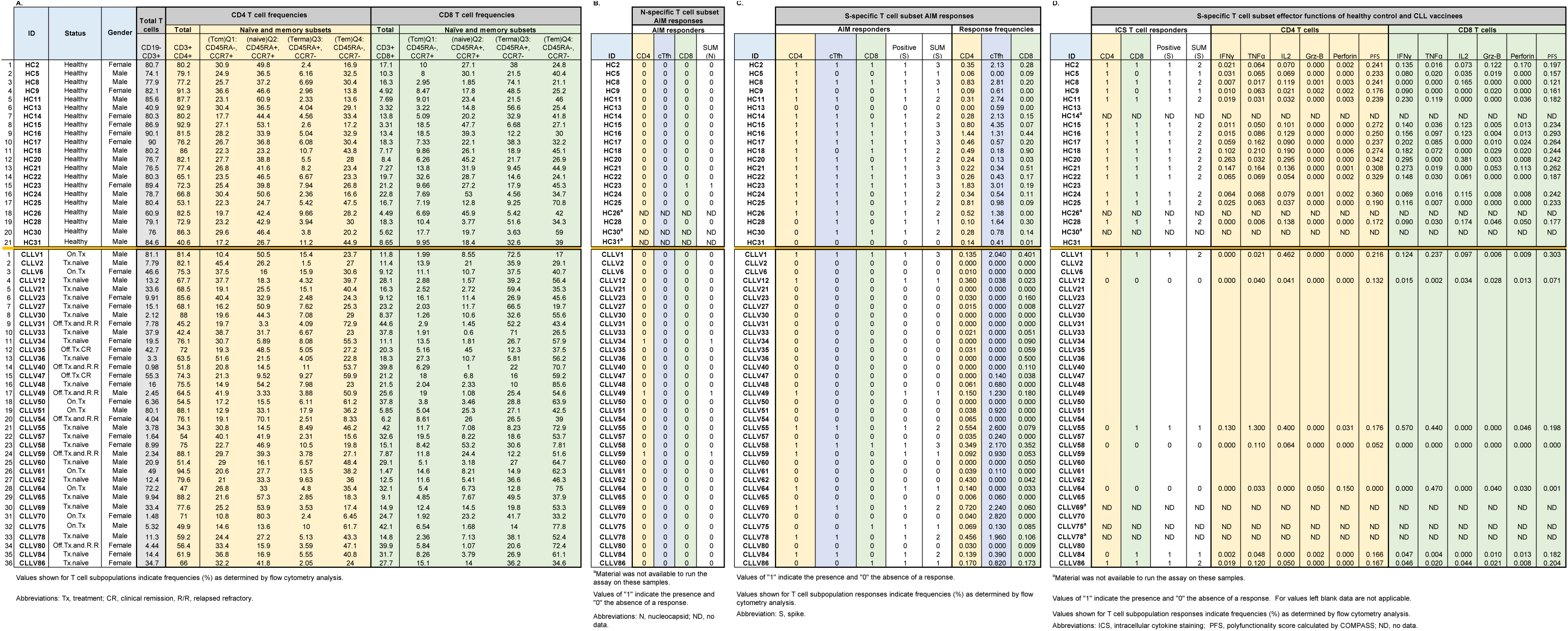
Subset frequencies, SARS-CoV-2 S peptide-specific AIM responses, and effector functions in healthy control and CLL donor T cells, Related to Figures 2, 5, S1 & S2.

## REFERENCES

1. SEER (2014-2018). Surveillance, Epidemiology, and End Results (SEER) Program. doi: https://seer.cancer.gov/statfacts/html/clyl.html.

2. Forconi, F., and Moss, P. (2015). Perturbation of the normal immune system in patients with CLL. Blood 126, 573–581. doi:10.1182/blood-2015-03-567388.

3. Hallek, M., Cheson, B.D., Catovsky, D., Caligaris-Cappio, F., Dighiero, G., Dohner, H., Hillmen, P., Keating, M., Montserrat, E., Chiorazzi, N. et al. (2018). iwCLL guidelines for diagnosis, indications for treatment, response assessment, and supportive management of CLL. Blood 131, 2745–2760. doi:10.1182/blood-2017-09-806398.

4. Sun, C., Gao, J., Couzens, L., Tian, X., Farooqui, M.Z., Eichelberger, M.C., and Wiestner, A. (2016). Seasonal Influenza Vaccination in Patients With Chronic Lymphocytic Leukemia Treated With Ibrutinib. JAMA Oncol 2, 1656–1657. doi:10.1001/jamaoncol.2016.2437.

5. Douglas, A.P., Trubiano, J.A., Barr, I., Leung, V., Slavin, M.A., and Tam, C.S. (2017). Ibrutinib may impair serological responses to influenza vaccination. Haematologica 102, e397–e399. doi:10.3324/haematol.2017.164285.

6. Svensson, T., Kattstrom, M., Hammarlund, Y., Roth, D., Andersson, P.O., Svensson, M., Nilsson, I., Rombo, L., Cherif, H., and Kimby, E. (2018). Pneumococcal conjugate vaccine triggers a better immune response than pneumococcal polysaccharide vaccine in patients with chronic lymphocytic leukemia A randomized study by the Swedish CLL group. Vaccine 36, 3701–3707. doi:10.1016/j.vaccine.2018.05.012.

7. Mauro, F.R., Giannarelli, D., Galluzzo, C.M., Vitale, C., Visentin, A., Riemma, C., Rosati, S., Porrazzo, M., Pepe, S., Coscia, M. et al. (2021). Response to the conjugate pneumococcal vaccine (PCV13) in patients with chronic lymphocytic leukemia (CLL). Leukemia 35, 737–746. doi:10.1038/s41375-020-0884-z.

8. Pleyer, C., Ali, M.A., Cohen, J.I., Tian, X., Soto, S., Ahn, I.E., Gaglione, E.M., Nierman, P., Marti, G.E., Hesdorffer, C. et al. (2021). Effect of Bruton tyrosine kinase inhibitor on efficacy of adjuvanted recombinant hepatitis B and zoster vaccines. Blood 137, 185–189. doi:10.1182/blood.2020008758.

9. Na, I.K., Buckland, M., Agostini, C., Edgar, J.D.M., Friman, V., Michallet, M., Sanchez-Ramon, S., Scheibenbogen, C., and Quinti, I. (2019). Current clinical practice and challenges in the management of secondary immunodeficiency in hematological malignancies. Eur. J. Haematol. 102, 447–456. doi:10.1111/ejh.13223.

10. Forni, D., Cagliani, R., Pozzoli, U., Mozzi, A., Arrigoni, F., De Gioia, L., Clerici, M., and Sironi, M. (2022). Dating the Emergence of Human Endemic Coronaviruses. Viruses 14. doi:10.3390/v14051095.

11. Edridge, A.W.D., Kaczorowska, J., Hoste, A.C.R., Bakker, M., Klein, M., Loens, K., Jebbink, M.F., Matser, A., Kinsella, C.M., Rueda, P. et al. (2020). Seasonal coronavirus protective immunity is short-lasting. Nat. Med. 26, 1691–1693. doi:10.1038/s41591-020-1083-1.

12. Dong, E., Du, H., and Gardner, L. (2020). An interactive web-based dashboard to track COVID-19 in real time. Lancet Infect. Dis. 20, 533–534. doi:10.1016/S1473-3099(20)30120-1.

13. Fung, M., and Babik, J.M. (2021). COVID-19 in Immunocompromised Hosts: What We Know So Far. Clin. Infect. Dis. 72, 340–350. doi:10.1093/cid/ciaa863.

14. Cascella, M., Rajnik, M., Aleem, A., Dulebohn, S.C., and Di Napoli, R. (2022). Features, Evaluation, and Treatment of Coronavirus (COVID-19). In StatPearls, (StatPearls Publishing Copyright © 2021, StatPearls Publishing LLC.).

15. Mato, A.R., Roeker, L.E., Lamanna, N., Allan, J.N., Leslie, L., Pagel, J.M., Patel, K., Osterborg, A., Wojenski, D., Kamdar, M. et al. (2020). Outcomes of COVID-19 in patients with CLL: a multicenter international experience. Blood 136, 1134–1143. doi:10.1182/blood.2020006965.

16. Chatzikonstantinou, T., Kapetanakis, A., Scarfo, L., Karakatsoulis, G., Allsup, D., Cabrero, A.A., Andres, M., Antic, D., Baile, M., Baliakas, P. et al. (2021). COVID-19 severity and mortality in patients with CLL: an update of the international ERIC and Campus CLL study. Leukemia 35, 3444–3454. doi:10.1038/s41375-021-01450-8.

17. Niemann, C.U., da Cunha-Bang, C., Helleberg, M., Ostrowski, S.R., and Brieghel, C. (2022). Patients with CLL have a lower risk of death from COVID-19 in the Omicron era. Blood 140, 445–450. doi:10.1182/blood.2022016147.

18. Walls, A.C., Park, Y.J., Tortorici, M.A., Wall, A., McGuire, A.T., and Veesler, D. (2020). Structure, Function, and Antigenicity of the SARS-CoV-2 Spike Glycoprotein. Cell 181, 281–292 e286. doi:10.1016/j.cell.2020.02.058.

19. Hoffmann, M., Kleine-Weber, H., Schroeder, S., Kruger, N., Herrler, T., Erichsen, S., Schiergens, T.S., Herrler, G., Wu, N.H., Nitsche, A. et al. (2020). SARS-CoV-2 Cell Entry Depends on ACE2 and TMPRSS2 and Is Blocked by a Clinically Proven Protease Inhibitor. Cell 181, 271–280 e278. doi:10.1016/j.cell.2020.02.052.

20. Avanzato, V.A., Matson, M.J., Seifert, S.N., Pryce, R., Williamson, B.N., Anzick, S.L., Barbian, K., Judson, S.D., Fischer, E.R., Martens, C. et al. (2020). Case Study: Prolonged Infectious SARS-CoV-2 Shedding from an Asymptomatic Immunocompromised Individual with Cancer. Cell 183, 1901–1912 e1909. doi:10.1016/j.cell.2020.10.049.

21. Jiang, S., Zhang, X., Yang, Y., Hotez, P.J., and Du, L. (2020). Neutralizing antibodies for the treatment of COVID-19. Nat Biomed Eng 4, 1134–1139. doi:10.1038/s41551-020-00660-2.

22. Honjo, K., Russell, R.M., Li, R., Liu, W., Stoltz, R., Tabengwa, E.M., Hua, Y., Prichard, L., Kornbrust, A.N., Sterrett, S. et al. (2021). Convalescent plasma-mediated resolution of COVID-19 in a patient with humoral immunodeficiency. Cell Rep Med 2, 100164. doi:10.1016/j.xcrm.2020.100164.

23. Gilbert, P.B., Montefiori, D.C., McDermott, A.B., Fong, Y., Benkeser, D., Deng, W., Zhou, H., Houchens, C.R., Martins, K., Jayashankar, L. et al. (2022). Immune correlates analysis of the mRNA-1273 COVID-19 vaccine efficacy clinical trial. Science 375, 43–50. doi:10.1126/science.abm3425.

24. Herishanu, Y., Avivi, I., Aharon, A., Shefer, G., Levi, S., Bronstein, Y., Morales, M., Ziv, T., Shorer Arbel, Y., Scarfo, L. et al. (2021). Efficacy of the BNT162b2 mRNA COVID-19 vaccine in patients with chronic lymphocytic leukemia. Blood 137, 3165–3173. doi:10.1182/blood.2021011568.

25. Greenberger, L.M., Saltzman, L.A., Senefeld, J.W., Johnson, P.W., DeGennaro, L.J., and Nichols, G.L. (2021). Antibody response to SARS-CoV-2 vaccines in patients with hematologic malignancies. Cancer Cell 39, 1031–1033. doi:10.1016/j.ccell.2021.07.012.

26. Blixt, L., Wullimann, D., Aleman, S., Lundin, J., Chen, P., Gao, Y., Cuapio, A., Akber, M., Lange, J., Rivera-Ballesteros, O. et al. (2022). T-cell immune responses following vaccination with mRNA BNT162b2 against SARS-CoV-2 in patients with chronic lymphocytic leukemia: results from a prospective open-label clinical trial. Haematologica 107, 1000–1003. doi:10.3324/haematol.2021.280300.

27. Liu, W., Russell, R.M., Bibollet-Ruche, F., Skelly, A.N., Sherrill-Mix, S., Freeman, D.A., Stoltz, R., Lindemuth, E., Lee, F.H., Sterrett, S. et al. (2021). Predictors of Nonseroconversion after SARS-CoV-2 Infection. Emerg. Infect. Dis. 27, 2454–2458. doi:10.3201/eid2709.211042.

28. Korber, B., Fischer, W.M., Gnanakaran, S., Yoon, H., Theiler, J., Abfalterer, W., Hengartner, N., Giorgi, E.E., Bhattacharya, T., Foley, B. et al. (2020). Tracking Changes in SARS-CoV-2 Spike: Evidence that D614G Increases Infectivity of the COVID-19 Virus. Cell 182, 812–827 e819. doi:10.1016/j.cell.2020.06.043.

29. Liu, C., Ginn, H.M., Dejnirattisai, W., Supasa, P., Wang, B., Tuekprakhon, A., Nutalai, R., Zhou, D., Mentzer, A.J., Zhao, Y. et al. (2021). Reduced neutralization of SARS-CoV-2 B.1.617 by vaccine and convalescent serum. Cell 184, 4220–4236 e4213. doi:10.1016/j.cell.2021.06.020.

30. Wang, L., Zhou, T., Zhang, Y., Yang, E.S., Schramm, C.A., Shi, W., Pegu, A., Oloniniyi, O.K., Henry, A.R., Darko, S. et al. (2021). Ultrapotent antibodies against diverse and highly transmissible SARS-CoV-2 variants. Science 373. doi:10.1126/science.abh1766.

31. Schmidt, F., Weisblum, Y., Muecksch, F., Hoffmann, H.H., Michailidis, E., Lorenzi, J.C.C., Mendoza, P., Rutkowska, M., Bednarski, E., Gaebler, C. et al. (2020). Measuring SARS-CoV-2 neutralizing antibody activity using pseudotyped and chimeric viruses. J. Exp. Med. 217. doi:10.1084/jem.20201181.

32. Parry, H., McIlroy, G., Bruton, R., Damery, S., Tyson, G., Logan, N., Davis, C., Willett, B., Zuo, J., Ali, M. et al. (2022). Impaired neutralisation of SARS-CoV-2 delta variant in vaccinated patients with B cell chronic lymphocytic leukaemia. J. Hematol. Oncol. 15, 3. doi:10.1186/s13045-021-01219-7.

33. Platsoucas, C.D., Galinski, M., Kempin, S., Reich, L., Clarkson, B., and Good, R.A. (1982). Abnormal T lymphocyte subpopulations in patients with B cell chronic lymphocytic leukemia: an analysis by monoclonal antibodies. J. Immunol. 129, 2305–2312.

34. Peller, S., and Kaufman, S. (1991). Decreased CD45RA T cells in B-cell chronic lymphatic leukemia patients: correlation with disease stage. Blood 78, 1569–1573.

35. Schreeder, D.M., Pan, J., Li, F.J., Vivier, E., and Davis, R.S. (2008). FCRL6 distinguishes mature cytotoxic lymphocytes and is upregulated in patients with B-cell chronic lymphocytic leukemia. Eur. J. Immunol. 38, 3159–3166. doi:10.1002/eji.200838516.

36. Riches, J.C., Davies, J.K., McClanahan, F., Fatah, R., Iqbal, S., Agrawal, S., Ramsay, A.G., and Gribben, J.G. (2013). T cells from CLL patients exhibit features of T-cell exhaustion but retain capacity for cytokine production. Blood 121, 1612–1621. doi:10.1182/blood-2012-09-457531.

37. Tarke, A., Sidney, J., Kidd, C.K., Dan, J.M., Ramirez, S.I., Yu, E.D., Mateus, J., da Silva Antunes, R., Moore, E., Rubiro, P. et al. (2021). Comprehensive analysis of T cell immunodominance and immunoprevalence of SARS-CoV-2 epitopes in COVID-19 cases. Cell Rep Med 2, 100204. doi:10.1016/j.xcrm.2021.100204.

38. Mateus, J., Grifoni, A., Tarke, A., Sidney, J., Ramirez, S.I., Dan, J.M., Burger, Z.C., Rawlings, S.A., Smith, D.M., Phillips, E. et al. (2020). Selective and cross-reactive SARS-CoV-2 T cell epitopes in unexposed humans. Science 370, 89–94. doi:10.1126/science.abd3871.

39. Lin, L., Finak, G., Ushey, K., Seshadri, C., Hawn, T.R., Frahm, N., Scriba, T.J., Mahomed, H., Hanekom, W., Bart, P.A. et al. (2015). COMPASS identifies T-cell subsets correlated with clinical outcomes. Nat. Biotechnol. 33, 610–616. doi:10.1038/nbt.3187.

40. Cyster, J.G., and Allen, C.D.C. (2019). B Cell Responses: Cell Interaction Dynamics and Decisions. Cell 177, 524–540. doi:10.1016/j.cell.2019.03.016.

41. Mesin, L., Ersching, J., and Victora, G.D. (2016). Germinal Center B Cell Dynamics. Immunity 45, 471–482. doi:10.1016/j.immuni.2016.09.001.

42. de Bourcy, C.F., Angel, C.J., Vollmers, C., Dekker, C.L., Davis, M.M., and Quake, S.R. (2017). Phylogenetic analysis of the human antibody repertoire reveals quantitative signatures of immune senescence and aging. Proc. Natl. Acad. Sci. U. S. A. 114, 1105–1110. doi:10.1073/pnas.1617959114.

43. Henry, C., Zheng, N.Y., Huang, M., Cabanov, A., Rojas, K.T., Kaur, K., Andrews, S.F., Palm, A.E., Chen, Y.Q., Li, Y. et al. (2019). Influenza Virus Vaccination Elicits Poorly Adapted B Cell Responses in Elderly Individuals. Cell Host Microbe 25, 357–366 e356. doi:10.1016/j.chom.2019.01.002.

44. Francis, T. (1960). On the doctrine of original antigenic sin. Proceedings of the American Philosophical Society 104, 572–578.

45. Whitaker, J.A., Parikh, S.A., Shanafelt, T.D., Kay, N.E., Kennedy, R.B., Grill, D.E., Goergen, K.M., Call, T.G., Kendarian, S.S., Ding, W. et al. (2021). The humoral immune response to high-dose influenza vaccine in persons with monoclonal B-cell lymphocytosis (MBL) and chronic lymphocytic leukemia (CLL). Vaccine 39, 1122–1130. doi:10.1016/j.vaccine.2021.01.001.

46. Herishanu, Y., Rahav, G., Levi, S., Braester, A., Itchaki, G., Bairey, O., Dally, N., Shvidel, L., Ziv-Baran, T., Polliack, A. et al. (2022). Efficacy of a third BNT162b2 mRNA COVID-19 vaccine dose in patients with CLL who failed standard 2-dose vaccination. Blood 139, 678–685. doi:10.1182/blood.2021014085.

47. Greenberger, L.M., Saltzman, L.A., Gruenbaum, L.M., Xu, J., Reddy, S.T., Senefeld, J.W., Johnson, P.W., Fields, P.A., Sanders, C., DeGennaro, L.J. et al. (2022). Anti-spike T-cell and Antibody Responses to SARS-CoV-2 mRNA Vaccines in Patients with Hematologic Malignancies. Blood Cancer Discov 3, 481–489. doi:10.1158/2643-3230.BCD-22-0077.

48. Farcet, M.R., Karbiener, M., Knotzer, S., Schwaiger, J., and Kreil, T.R. (2022). Omicron Severe Acute Respiratory Syndrome Coronavirus 2 Neutralization by Immunoglobulin Preparations Manufactured From Plasma Collected in the United States and Europe. J. Infect. Dis. 226, 1396–1400. doi:10.1093/infdis/jiac358.

49. Bruel, T., Hadjadj, J., Maes, P., Planas, D., Seve, A., Staropoli, I., Guivel-Benhassine, F., Porrot, F., Bolland, W.H., Nguyen, Y. et al. (2022). Serum neutralization of SARS-CoV-2 Omicron sublineages BA.1 and BA.2 in patients receiving monoclonal antibodies. Nat. Med. 28, 1297–1302. doi:10.1038/s41591-022-01792-5.

50. Sacco, M.D., Hu, Y., Gongora, M.V., Meilleur, F., Kemp, M.T., Zhang, X., Wang, J., and Chen, Y. (2022). The P132H mutation in the main protease of Omicron SARS-CoV-2 decreases thermal stability without compromising catalysis or small-molecule drug inhibition. Cell Res. 32, 498–500. doi:10.1038/s41422-022-00640-y.

51. G Doukas, P., St Pierre, F., Boyer, J., Nieves, M., and Ma, S. (2022). CLO22-043: Humoral Immune Response Following COVID-19 Vaccination in Patients With Chronic Lymphocytic Leukemia and Other Indolent Lymphomas: A Large, Single-Center Observational Study. J. Natl. Compr. Canc. Netw. 20, CLO22-043–CLO022-043. doi:10.6004/jnccn.2021.7248.

52. Mitchell, J., Connolly, C.M., Chiang, T.P., Alejo, J.L., Werbel, W.A., Segev, D.L., and Massie, A.B. (2022). Comparison of SARS-CoV-2 Antibody Response After 2-Dose mRNA-1273 vs BNT162b2 Vaccines in Incrementally Immunosuppressed Patients. JAMA Netw Open 5, e2211897. doi:10.1001/jamanetworkopen.2022.11897.

53. Baden, L.R., El Sahly, H.M., Essink, B., Kotloff, K., Frey, S., Novak, R., Diemert, D., Spector, S.A., Rouphael, N., Creech, C.B. et al. (2021). Efficacy and Safety of the mRNA-1273 SARS-CoV-2 Vaccine. N. Engl. J. Med. 384, 403–416. doi:10.1056/NEJMoa2035389.

54. Polack, F.P., Thomas, S.J., Kitchin, N., Absalon, J., Gurtman, A., Lockhart, S., Perez, J.L., Perez Marc, G., Moreira, E.D., Zerbini, C. et al. (2020). Safety and Efficacy of the BNT162b2 mRNA Covid-19 Vaccine. N. Engl. J. Med. 383, 2603–2615. doi:10.1056/NEJMoa2034577.

55. Anderson, E.J., Rouphael, N.G., Widge, A.T., Jackson, L.A., Roberts, P.C., Makhene, M., Chappell, J.D., Denison, M.R., Stevens, L.J., Pruijssers, A.J. et al. (2020). Safety and Immunogenicity of SARS-CoV-2 mRNA-1273 Vaccine in Older Adults. N. Engl. J. Med. 383, 2427–2438. doi:10.1056/NEJMoa2028436.

56. Li, Y., Tenchov, R., Smoot, J., Liu, C., Watkins, S., and Zhou, Q. (2021). A Comprehensive Review of the Global Efforts on COVID-19 Vaccine Development. ACS Cent Sci 7, 512–533. doi:10.1021/acscentsci.1c00120.

57. Boppana, S., Qin, K., Files, J.K., Russell, R.M., Stoltz, R., Bibollet-Ruche, F., Bansal, A., Erdmann, N., Hahn, B.H., and Goepfert, P.A. (2021). SARS-CoV-2-specific circulating T follicular helper cells correlate with neutralizing antibodies and increase during early convalescence. PLoS Pathog. 17, e1009761. doi:10.1371/journal.ppat.1009761.

58. Bryan, A., Pepper, G., Wener, M.H., Fink, S.L., Morishima, C., Chaudhary, A., Jerome, K.R., Mathias, P.C., and Greninger, A.L. (2020). Performance Characteristics of the Abbott Architect SARS-CoV-2 IgG Assay and Seroprevalence in Boise, Idaho. J. Clin. Microbiol. 58, JCM.00941-00920. doi:10.1128/JCM.00941-20.

59. Dohner, H., Stilgenbauer, S., Benner, A., Leupolt, E., Krober, A., Bullinger, L., Dohner, K., Bentz, M., and Lichter, P. (2000). Genomic aberrations and survival in chronic lymphocytic leukemia. N. Engl. J. Med. 343, 1910–1916. doi:10.1056/NEJM200012283432602.

60. Brouwer, P.J.M., Caniels, T.G., van der Straten, K., Snitselaar, J.L., Aldon, Y., Bangaru, S., Torres, J.L., Okba, N.M.A., Claireaux, M., Kerster, G. et al. (2020). Potent neutralizing antibodies from COVID-19 patients define multiple targets of vulnerability. Science 369, 643–650. doi:10.1126/science.abc5902.

61. Ketas, T.J., Chaturbhuj, D., Portillo, V.M.C., Francomano, E., Golden, E., Chandrasekhar, S., Debnath, G., Diaz-Tapia, R., Yasmeen, A., Kramer, K.D. et al. (2021). Antibody Responses to SARS-CoV-2 mRNA Vaccines Are Detectable in Saliva. Pathog Immun 6, 116–134. doi:10.20411/pai.v6i1.441.

62. Kumar, G., Sterrett, S., Hall, L., Tabengwa, E., Honjo, K., Larimer, M., Davis, R.S., Goepfert, P.A., and Larimer, B.M. (2022). Comprehensive mapping of SARS-CoV-2 peptide epitopes for development of a highly sensitive serological test for total and neutralizing antibodies. Protein Eng Des Sel 35, gzab033. doi:10.1093/protein/gzab033.

